# Cost-Sharing in Medical Care Can Increase Adult Mortality Risk in Lower-Income Countries

**DOI:** 10.1101/2021.03.03.21252857

**Authors:** Giancarlo Buitrago, Grant Miller, Marcos Vera-Hernández

**Affiliations:** Instituto de Investigaciones Clínicas, Facultad de Medicina, Universidad Nacional de Colombia; and Hospital Universitario Nacional de Colombia, Bogotá, Colombia; Department of Medicine, Stanford University, Stanford, CA; and National Bureau of Economic Research, Cambridge, MA, USA; Department of Economics, University College London; and Institute for Fiscal Studies, London, UK

## Abstract

Patient cost-sharing in medical care constrains total health spending, presumably with little harm to underlying patient health. This paper re-evaluates the link between cost-sharing and health, studying Colombia’s entire formal sector workforce with individual-level health care utilization records linked to payroll data and vital statistics. Given discrete breaks in outpatient cost-sharing imposed at multiple income thresholds by Colombia’s national health system, we use a regression discontinuity design and find that outpatient cost-sharing reduces use of outpatient care, resulting in fewer diagnoses of common chronic diseases and increasing subsequent emergency room visits and hospitalizations. Ultimately, these effects measurably increase mortality, and disproportionately so among the poor – raising the absolute difference in 7-year mortality risk by 0.80 and 0.23 deaths per 1,000 individuals at lower- and higher-income thresholds, respectively. To the best of our knowledge, this study is the first to show a relationship between cost-sharing and adult mortality risk in lower-income countries, a relationship important to incorporate into social welfare analyses of cost-sharing policies.

**One Sentence Summary:** Outpatient cost-sharing in medical care discourages use – but over time, also increases costly hospital service use and raises mortality risk.

## Main Text

Consumer cost-sharing in medical care (through co-payments, co-insurance, and deductibles) is strongly related to the use of health care services and spending – but it has never been shown to influence adult mortality risk in lower-income countries.^1^ Traditionally, the role of cost-sharing under health insurance is to balance protection against financial risk with overuse of medical care (i.e., moral hazard) (Arrow 1963; Pauly 1968; Zeckhauser 1970)– and to constrain total health care spending (Manning et al. 1987; Newhouse and Insurance Experiment Group 1996; Chernew and Newhouse 2008). However, cost sharing can also be associated with reductions in preventive care, disease detection, and the use of clinically important services, potentially leading to costly increases in hospital care (Brot-Goldberg et al. 2017; Chandra, Gruber, and McKnight 2010). This concern may be particularly true for prevalent chronic (rather than acute) conditions such as hypertension and diabetes which often develop and progress undetected in their early stages without regular clinical consultation and monitoring. Chronic disease prevalence is rising in all parts of the world, in wealthy and poor countries alike, accounting for an increasingly large share of the global burden of disease and deaths (Ezzati and Riboli 2012; Gaziano and Pagidipati 2013; NCD Countdown 2030 Collaborators 2018).^2^

To the best of our knowledge, this study is the first population-level analysis to present evidence showing a causal relationship between cost-sharing and adult mortality risk in a lower-income country.^3^ We study the universe of Colombia’s formal sector workforce over the span of nearly a decade using monthly health care claims data linked at the individual level to administrative payroll data and vital statistics. With this data, we take advantage of discrete changes in outpatient co-payment requirements at well-defined wage rate thresholds using a regression-discontinuity study design. Conditional on the assumption that wage rates are not manipulated or ‘gamed’ to obtain eligibility for lower copayments (an assumption for which we show empirical support), regression-discontinuity designs are able to provide internally valid estimates of causal relationships (Thistlethwaite and Campbell 1960; Hahn, Todd, and Van der Klaauw 2001; Lee and Lemieux 2010; Bor et al. 2014). Moreover, given the institutional design of Colombia’s health care system, we are able to conduct this analysis at two different wage/income levels, enabling us to assess the effect of cost-sharing at mortality risk at different points in the income distribution.

The outpatient cost-sharing hikes imposed at discrete breaks in the wage distribution can be substantial, especially for poorer households. Colombia’s sweeping health insurance reform in 1993 introduced competition among health plans into the Colombian health care system (through a form of managed competition financed by payroll taxes), but both benefits and prices (premiums – and importantly, patient cost-sharing) are fully standardized across all competing insurers. In the Contributory Regime, or *Régimen Contributivo* (for all formal sector employees and their families – numbering approximately 20 million enrollees), there are discrete outpatient cost-sharing “steps” at two different thresholds in the distribution of monthly earnings. Specifically, for outpatient services, individuals earning less than twice Colombia’s monthly minimum wage (MMW) must pay 11.7% of the daily minimum wage per outpatient service, those earning two up to five times (inclusive) of the MMW pay 46.1% of the daily minimum wage, and those earning above five MMWs pay 121.5% of the daily minimum wage. **Supplement Figure S1** shows these outpatient cost-sharing requirements across the earnings distribution. Note that there is no cost-sharing requirement for inpatient services. Classification of earnings is maintained by the Colombian Ministry of Finance and its Financial Services Authority in a digital system known as *Planilla Integrada de Liquidación de Aportes*, or PILA, and all formal sector companies update this information monthly as part of their mandated payroll tax contributions to the Colombian social security system.

Our data are novel and represents an important collaboration with Colombian national government agencies to provide a stronger evidence-base for health policy. Specifically, we link individual records for nearly 13 million Colombians across: (i.) monthly data on payroll and tax contributions (from the *Planilla Integrada de Liquidación de Aportes*, which is part of the PILA system); (ii.) annual health insurance enrollment (in the *Base Única de Afiliación*, or BDUA); (iii.) monthly health service use, claims, and diagnosis data (in the *Base del Estudio de la Suficiencia de la Unidad por Capitación*, or UPC; **Supplement Table S1** shows ICD10 codes for chronic diseases); and (iv.) national vital statistics. **Supplement Table S2** shows descriptive statistics for this linked administrative dataset.

We first use a regression discontinuity design (RDD) to estimate the causal relationship between cost-sharing requirements and outpatient service use. Specifically, we use a standard local linear regression in which the outcome variable, *y*_*imt*_, refers to outpatient services for individual *i* in month *m* and year *t*, and the running variable is the individual’s *i* monthly earnings as a share of the monthly minimum wage. We estimate two separate regressions, one for each threshold of the earnings distribution (2 or 5 MMWs). We select robust bias-corrected ‘optimal’ sample bandwidths, and we adjust standard errors for heteroskedasticity and clustering at the individual level (Calonico, Cattaneo, and Titiunik 2014; Calonico, Cattaneo, and Farrell 2020).

In this statistical framework, we assume that monthly earnings registered by formal-sector companies in the PILA system are not manipulated or ‘gamed’ to obtain eligibility for lower copayments. If individuals were able manipulate their earnings (or how their earnings are registered in the PILA system) to obtain a lower outpatient copayment rate, this would be evident as imbalance along demographic dimensions related to their ability or incentive to do so. To test for this, **Supplement Figures S2 and S3** show balance, or the absence of a discontinuity, in demographic characteristics (including age and gender as well as enrollment in the public plan offered through Colombia’s managed competition style of health insurance) across both cost-sharing thresholds. Then, to test for statistical significance we randomly draw 500 false thresholds and estimate the local linear regressions using the demographic characteristics as dependent variable. **Supplement Figure S4** shows that the null hypothesis of continuity in these variables is not rejected, implying the absence of manipulation. We also implement an alternative test for manipulation of earnings, formally testing for disproportionate mass in the earnings distribution on either side of each threshold (following (McCrary 2008)). If individuals manipulated their reported earnings, and hence their copayment tier, this would be evidence as disproportionate mass just below a threshold. **Supplement Figure S5** shows these results, again providing no evidence of manipulation (McCrary tests: 0.15 (p-value 0.21) and 0.06 (p-value 0.42) for thresholds at 2 and 5 MMWs, respectively).

Examining the direct effect of outpatient cost-sharing on the use of outpatient services, **Figure 1** shows discrete reductions in total outpatient services used at both the 2 and 5 MMW thresholds (average reductions of 0.12 and 0.06 services per month, or reductions of 12.90% and 8.70%, respectively). Breaking this cost-sharing effect into its components (consultations, drugs, lab procedures, and diagnostic imaging, which together sum to our total outpatient service measure) and considering annual (rather than monthly) outpatient care use, **Figure 2** shows separate results for component.^4^ In general, outpatient care reductions are largely due to decreases in drug purchases and clinical consultations (the components most under patient control). Strikingly, both figures show that cost-sharing reduces outpatient care relatively more for the poor (i.e., the reduction is larger at the 2 relatives to the 5 minimum wage threshold), but the magnitude of the cost-sharing change also differs at the two thresholds. To make a direct comparison between the two, **Supplement Tables S5 and S6** also report price elasticities at both thresholds – confirming that the decline is indeed larger for poorer households.^5^

**Fig. 1.**
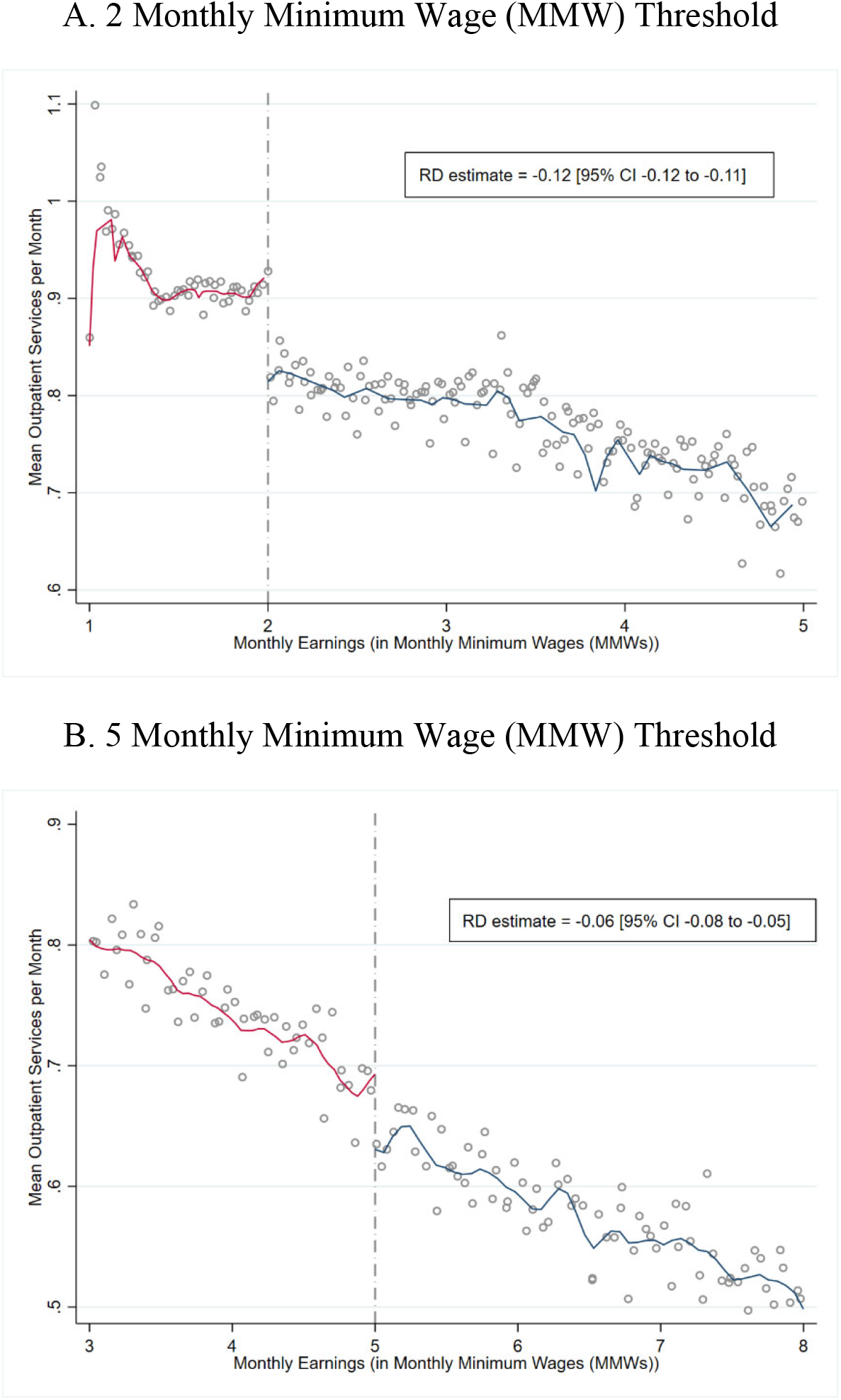
Total Outpatient Service Use across the Copayment Threshold. Regression discontinuity (RD) graphs with local linear smoothing for both sides of the threshold at 2 monthly minimum wages **(A)** and 5 monthly minimum wages **(B)**. Vertical axis show means of total number of outpatient services used per month for all formal workers between 2011 and 2017. We use local linear regression with robust bias-corrected ‘optimal’ sample bandwidths, standard errors adjusting for heteroskedasticity and clustering at the individual level, to estimate the effect of cost-sharing on total outpatient service use.

**Fig. 2.**
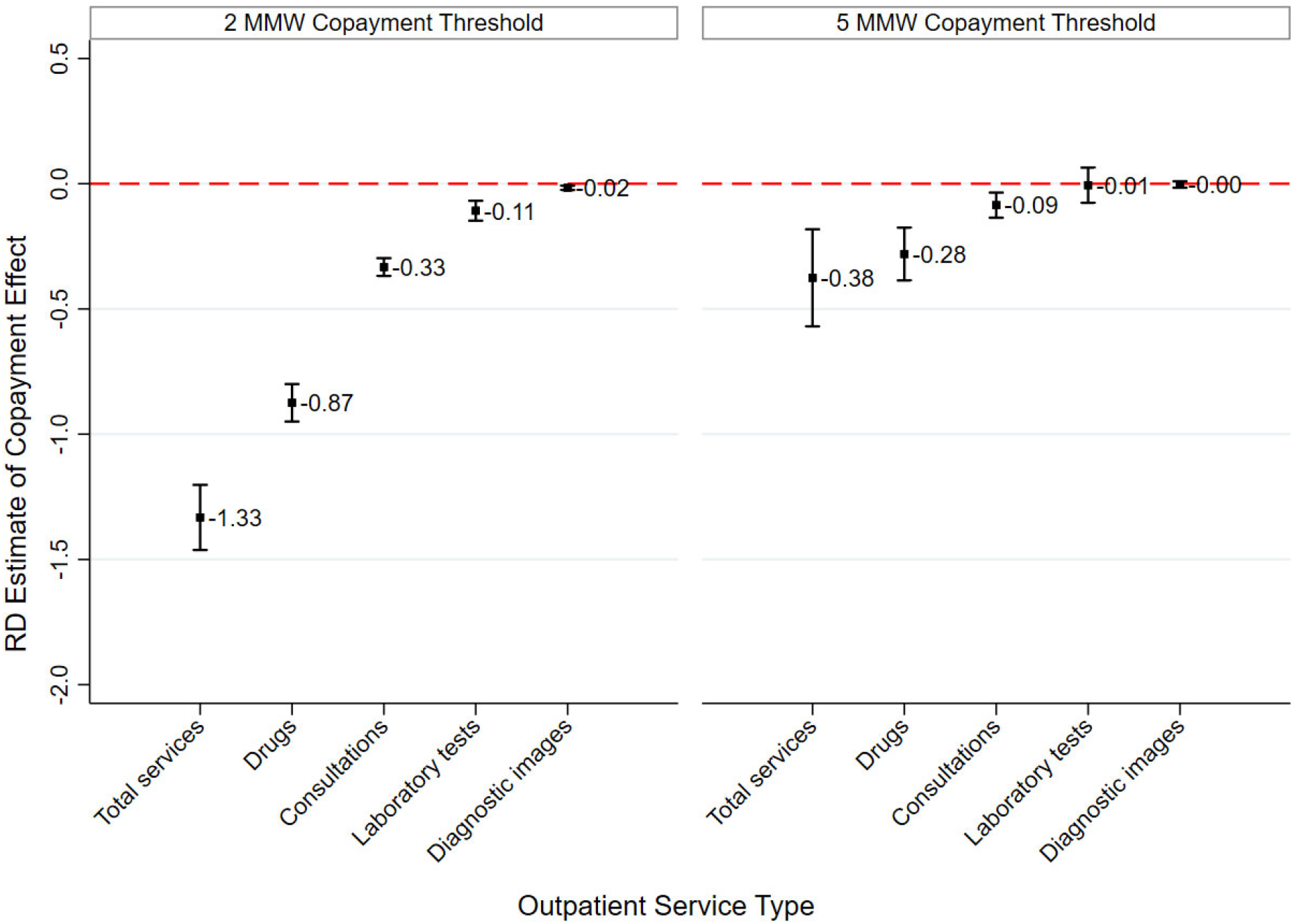
The Effect of Cost-Sharing on Outpatient Service Use, Total and by Type. Effect of greater outpatient cost-sharing on the annual use of total number of outpatient services and its components (consultations, drugs, laboratory procedures, and diagnostic images) for both thresholds (2 MMW and 5 MMW). We use a sample of individuals with median monthly earnings within 0.3 MMWs of the threshold, and who worked during the entire year (12 months). We use ordinary least square with a second-order polynomial of the monthly earnings, and we compute heteroskedastic standard errors clustered at the individual level.

Next, we extend our statistical framework to study how these ‘contemporaneous’ changes in outpatient service use accumulate over time to influence the subsequent detection and diagnosis of chronic diseases (including hypertension and diabetes, which are relatively prevalent in Colombia) – and as a result, the use of potentially avoidable inpatient and other hospital services. Specifically, we use OLS to estimate: 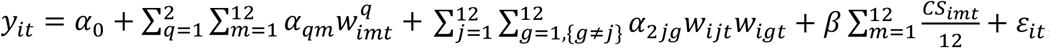, where the outcome variable *yit* (diagnosis of a chronic condition, hospitalization as an inpatient, emergency room (ER) use, or use of a hospital intensive care unit) for individual *i* in year *t*; *α*_*qm*_ and *α*_*2jg*_ multiply the polynomials of individual’s *i* twelve monthly wages (as share of the minimum wage) in year *t*; *CS*_*imt*_ takes value 0 or 1 (denoting below or above a threshold, respectively), and so 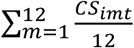 is the average number of months that individual *i* in year *t* is above a threshold; and *ε*_*it*_ is the error term. The parameter *β* measures the casual effect of cost sharing on diagnoses and inpatient (and other hospital) service use. We use a sample of individuals with median monthly earnings within 0.3 or 0.5 MMWs of the threshold (and who worked during the entire year), include a second-order wage polynomial (given evidence that higher-order polynomials can lead to biased results) (Gelman and Imbens 2019), and compute heteroskedastic standard errors clustered at the individual level.^6^

**Figure 3, Panel A** and **Supplement Tables S7-8** show these cumulative effects of cost-sharing for diagnoses (estimates of the parameter *β*), both for the same year and for all 7 years in the sample. Outpatient cost sharing leads to a reduced probability of chronic disease diagnosis, and these reductions are relatively larger for poorer households (at the 2 MMWs threshold relative to the 5 MMWs threshold). Moreover, the cumulative effect of cost-sharing on chronic disease diagnosis grows over time for poorer households as well. **Figure 3, Panel B** and **Supplement Tables S9-12** then show that greater cost sharing also subsequently leads to more use of costly, potentially avoidable hospital services – including inpatient stays, ER visits, and use of intensive care units.

**Fig. 3.**
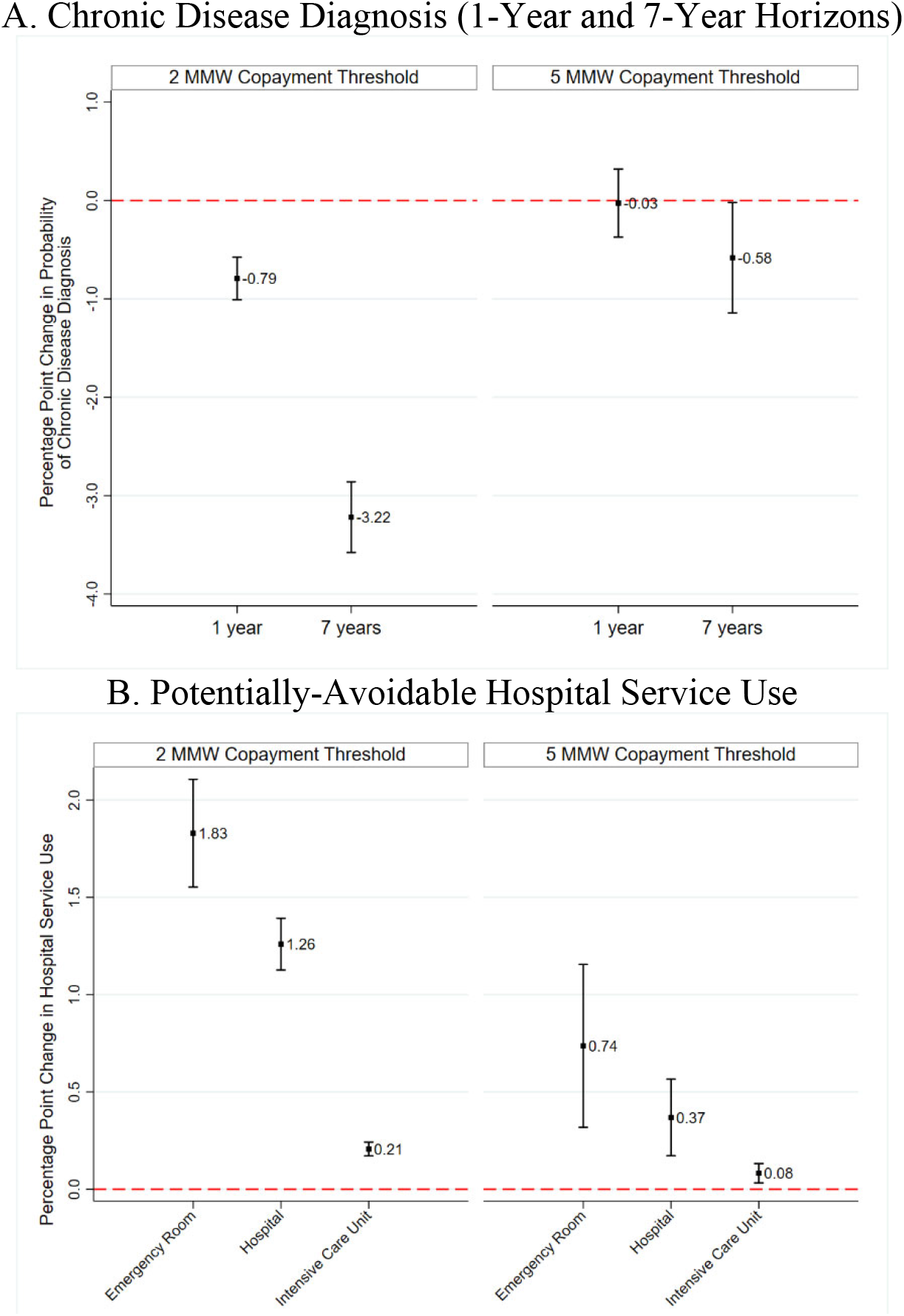
Cumulative Effect of Outpatient Cost-Sharing on Chronic Disease Diagnosis and Potentially-Avoidable Hospital Service Use. **A**. Effect of greater outpatient cost-sharing on the probabilities (percentage points) of detecting chronic diseases for the same year and for all 7 years in the sample at both thresholds (2 MMW and 5 MMW). **B**. Effect of greater outpatient cost-sharing on the probabilities (percentage points) of use of potentially avoidable hospital services – including inpatient stays, emergency room visits, and use of intensive care units, at both thresholds (2 MMW and 5 MMW). For all analysis, we use a sample of individuals with median monthly earnings within 0.3 MMWs of the threshold, and who worked during the entire year (12 months). We use ordinary least square with a second-order polynomial of the monthly earnings, and we compute heteroskedastic standard errors clustered at the individual level.

Finally, although the actual productivity of medical care services for health outcomes is the subject of ongoing debate, we examine how these changes in health service use due to cost-sharing matter for mortality (or survival). On the one hand, at lower levels of service use, additional services may be incrementally more productive (i.e., on the margin) simply because they are otherwise used at low rates. In lower-income countries, a variety of medical care services are in fact generally used less than in wealthy countries (Kruk et al. 2018; O’Donnell 2007). On the other hand, health care providers in lower-income countries might have fewer resources at their disposal (including health education specialists, etc.), suggesting that the incremental return to medical care may be low (particularly for illnesses requiring considerable lifestyle modification and private self-maintenance – as is the case for chronic conditions such as hypertension and diabetes, for example) (Kruk et al. 2018; Das and Hammer 2014).

To consider this issue directly, we estimate the cumulative effect of differences in service use due to outpatient cost-sharing (and changes in subsequent chronic disease detection as well as hospital and ER service use) by integrating our RDD into a duration analysis framework (Bor et al. 2014). This approach uses a semiparametric regression model to specify the mortality hazard (i.e., the instantaneous probability of death at time t, conditional on survival up to time t) as a function of the ‘running variable’ (in our case, the monthly earnings) and time. Because the monthly earnings vary over time, we use a time-dependent Cox hazards model, which is appropriate with ‘external’ covariates (i.e., covariates vary as a function of time, independent of the failure time) (Cox 1972; Prentice and Kalbfleisch 1979; Cox 1975; Kalbfleisch and Prentice 2011), and we explore the sensitivity of our results using other parametric assumptions (including Weibull and Exponential functions) as well as sample restrictions to those consistently employed in the formal sector over time. The Supplementary Materials describe these approaches in detail.

Figure 4. shows these differences in survival due to outpatient cost-sharing. **Figure 4, Panel A** shows 7-year survival curves estimated for each copayment tier (at both copayment thresholds) using time-dependent Cox models. It suggests that greater outpatient cost-sharing decreases 7-year survival at both thresholds. **Figure 4, Panel B** then shows corresponding hazard ratio estimates (to higher vs. lower copayment requirements) at both thresholds among samples of individuals continuously employed in the formal sector for varying amounts of time. It confirms that these reductions in survival are statistically significant (hazard ratios of 4.42 and 1.30 at the 2 and 5 MMW thresholds, respectively) and increase with the amount of time that individuals are subject to copayment requirements. Importantly, these effects on survival or longevity are also larger for individuals with lower earnings (i.e., at the 2 vs. 5 MMW threshold) – the corresponding elasticities are 1.06 and 0.29, implying increases in 7-year mortality risk by 0.80 and 0.23 deaths per 1,000 individuals at the 2 and 5 MMW thresholds, respectively. **Supplemental Tables S13** also shows these results in table format, and **Supplemental Tables S14-S15** show that these results are similar using Exponential and Weibull distributions.

**Fig. 4.**
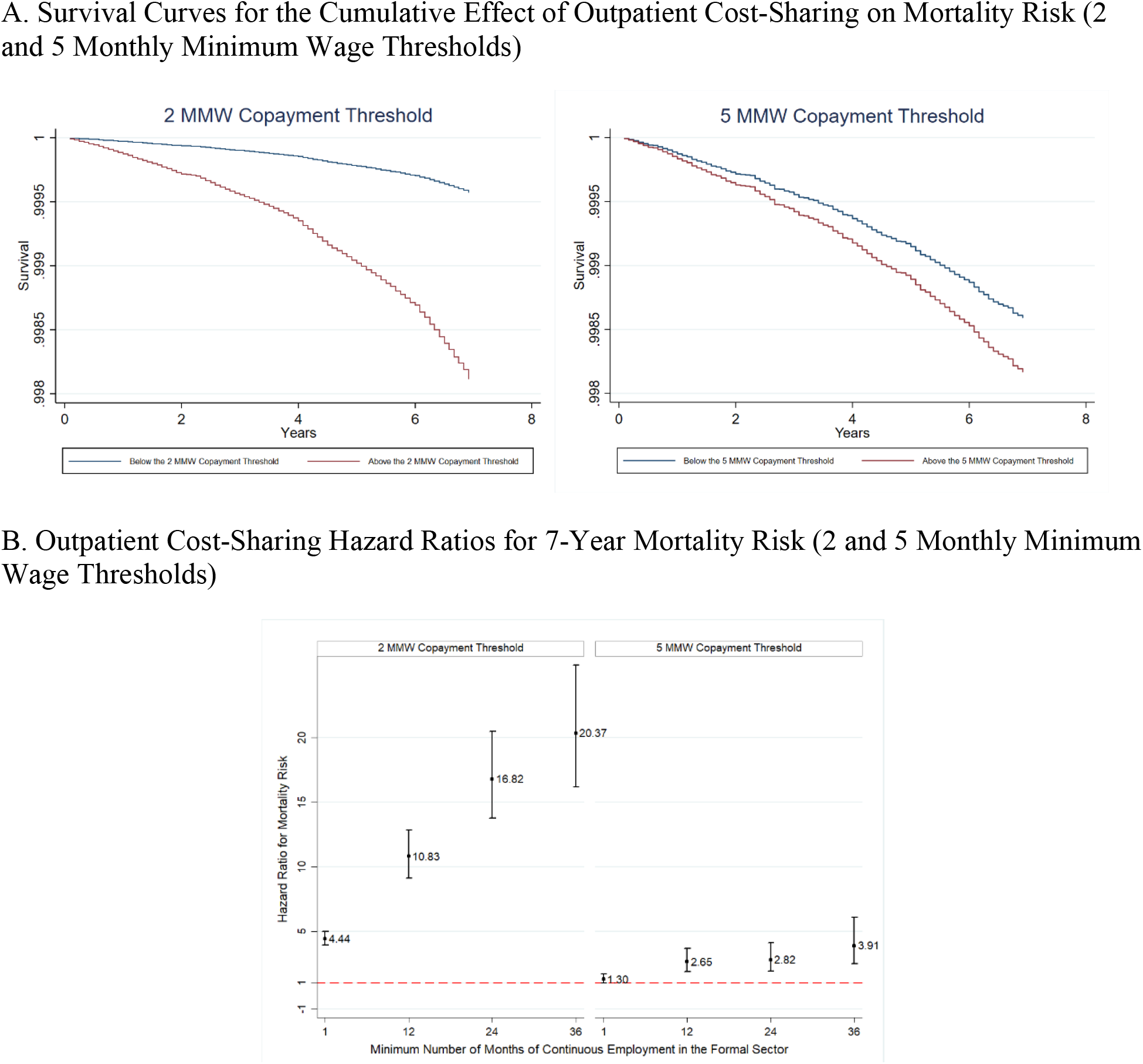
Cumulative Effect of Outpatient Cost-Sharing on 7-Year Survival. **A**. Survival curves for the cumulative effect of outpatient cost-sharing on mortality risk at both thresholds (2 MMW and 5 MMW) using time-dependent Cox models with full sample. **B**. Outpatient cost-sharing hazard ratios for 7-year mortality risk among samples of individuals continuously employed in the formal sector for varying amounts of time, and with median monthly earnings within 1 MMW of the thresholds. We use time-dependent Cox models with a second-order polynomial of the monthly earnings.

Overall, in this paper we provide new evidence that outpatient cost-sharing reduces the use of outpatient services – and in doing so, can unintentionally reduce the detection of new chronic diseases and increase the use of more expensive, and avoidable, hospital and ER services – ultimately increasing adult mortality. Although past research has traditionally emphasized the role of consumer cost-sharing in reducing potentially socially wasteful health care spending, this role must be balanced with considerations of detrimental effects of cost-sharing on health and mortality. This is an important direction for future social welfare evaluations of cost-sharing policies.

## Data Availability

The data were provided by the Ministry of Health and Social Protection of Colombia to the National University of Colombia. To access these data, requests should be submitted to the Ministry of Health and Social Protection of Colombia.

## Acknowledgments

We thank the Office of Information and Communication Technology of the Colombian Ministry of Health and Social Protection for providing the anonymized data for this study.

## Author Contributions

Buitrago: Conceptualization, Methodology, Writing, Funding Acquisition, Software, Formal Analysis

Miller: Conceptualization, Methodology, Writing, Supervision

Vera-Hernández: Conceptualization, Methodology, Writing, Funding Acquisition

## Data and Materials Availability

The data were provided by the Ministry of Health and Social Protection of Colombia to the National University of Colombia. To access these data, requests should be submitted to the Ministry of Health and Social Protection of Colombia. The code and log/output files for all analyses reported are publicly available and archived through the journal’s digital platform.

## Supplementary Materials

### 1. Materials and Methods

#### 1.1. Institutional Background

The current Colombian health care system (called *Sistema General de Seguridad Social en Salud*) was created in 1993 under Law 100. This social health insurance system offers a benefits package defined by Ministry of Health and administered by both public and private insurers. There are two major ‘regimes’ within this system: the ‘Contributory Regime’ and the ‘Subsidized Regime’. The Contributory Regime includes all formal-sector workers (and dependents) earning one or more legally-established monthly minimum wages (MMW). Alternatively, the Subsidized Regime covers all individuals (and dependents) earning less than one MMW and also meeting a proxy means test through the *Sistema de Identificación de Beneficiarios* (SISBEN). The benefits package is the same for both regimes and is generally comprehensive, covering all outpatient and inpatient services for almost all diseases, only some health technologies are excluded due to the absence of a sanitary register or non-clinical purposes (cosmetic plastic surgery, for example). Nearly the entire Colombian population is enrolled in one of these two regimes – in 2016, for example, the overall population coverage rate was 95.6%, with 45.54% in the Contributory Regime and 45.48% in the Subsidized Regime (Ministerio de Salud y Protección Social 2017).

Contributory Regime enrollees (called *Cotizantes*) face a step-function copayment for outpatient services (including consultations with general practitioners and specialists, drugs, and diagnostic tests) that varies with monthly earnings (measured in MMWs) and is officially recorded by Ministry of Finance using payroll data from employers. Specifically, there are three copayment tiers:

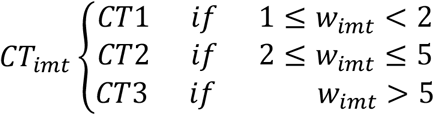

where *CT*_*imt*_ is the individual’s *i* copayment amount (in Colombian Pesos) in month *m* and year *t*; *w*_*imt*_ is individual’s *i* earnings (in monthly minimum wages units) in month *m* and year *t.^1^*

The corresponding copayment amount in each tier is:

*CT1*: 11.7% of a daily minimum wage, which roughly corresponds to 3,400 Colombian Pesos (COP $) (or 0.92 United States Dollars (USD $)) in 2020.

*CT2*: 46.1% of a daily minimum wage, which roughly corresponds to COP $ 13,500 (USD $ 3.65) in 2020.

*CT3*: 121.5% of a daily minimum wage, which roughly corresponds to COP $ 35,600 (USD $ 9.62) in 2020.

The copayment is paid by the worker (i.e., the *Cotizante*) for the outpatient care services use, which includes consultations with general practitioners and specialists, drugs, and diagnostic tests. There is no limit on the annual copayment that a worker can pay in a year.

Additionally, some outpatient services have no copayment requirement – most relevant to this study are those related to chronic disease management (for hypertension and diabetes, for example). There is no cost-sharing requirement for inpatient care.

#### 1.2. Data and Study Population

Our study includes all individuals enrolled in Contributory Regime for at least one month between January 2011 and December 2017. We excluded individuals who reached the legal retirement age (57 for women and 62 for men) in 2011 because the benefits to these individuals change as pensioners, but we are unable to identify pensioners from those still in the labor force.

To build our database of all Contributory Regime enrollees, we used the following data sources:

1. ‘Unique Affiliation Database’ (*Base de Datos Única de Afiliación*, or BDUA). The BDUA is the official government registrar tool for tracking and designating individual enrollee status in the Colombian health system. This database also includes basic socio-demographic characteristics of enrollees.
2. ‘Integrated Contribution Settlement Worksheet’ (*Planilla Integrada de Liquidación de Aportes*, or PILA). The PILA contains monthly payroll data on the economic contributions of citizens and their employers to Colombian social security systems, as reported by employers.
3. ‘Study Basis for Calculation of the Capitation Unit’ (*Base del Estudio de Suficiencia de la Unidad Por Capitación*, or UPC). The UPC database contains detailed records of each health service use by each Colombian enrolled in the country’s health care system (including identity of the enrollee, location of service, date of service, specific type of service, any diagnostic information, identity (and type) of health professional providing the service, and payments/reimbursements for the service). The UPC is the database used by the Ministry of Health and Social Protection for computation of risk-adjustment payments added to the insurance premiums paid to insurers.
4. Vital Statistics (*Registro Único de Afiliación*, or RUAF): The RUAF contains individual-level death certificate data maintained by the Colombian National Department of Statistics, including date of death, cause(s) of death, and geographic location of death.

Using these data sources, we first used PILA to identify all formal sector employees with incomes greater than or equal to one MMW in any month of the 84-month study period (January 2011-December 2017). Next, at the individual level, we link each person with the individual’s information in the BDUA database to merge health insurance enrollment status and socio-demographic characteristics. Then, using UPC data, we link each individual in the database with her health care utilization records for each service in each study month. Finally, we use RUAF data to identify each individual in our database who died during the study period, merging that individual with information about her death (death date, location, and cause(s)).

#### 1.3. Treatment Assignment

The primary exposure or treatment that we study is the copayment level that each individual Contributory Regime enrollee faced in each study month. We assign this exposure/treatment using the precise earnings (in MMW units) reported by employers to PILA.

#### 1.4. Outcomes

The primary outcome of our study is individuals’ survival time (time to death). The maximum survival time observed is seven years (84 months).

Additionally, we also study other primary outcomes that contribute to survival time: outpatient service use, inpatient service use, and new chronic disease diagnoses. Specifically:

##### Outpatient Services

1. OUT: Number of outpatient services during the period.
2. DRUGS: Number of drugs purchased during the period.
3. CONS: Number of medical consultations during the period.
4. LAB: Number of laboratory procedures during the period.
5. IMAG: Number of diagnostic imaging procedures during the period.

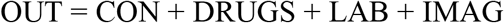

##### Inpatient Services

1. ER: An indicator variable taking value 1 if an individual visits the Emergency Room (ER) during the period, and 0 otherwise.
2. NUMER: Number ER visits during the period.
3. HOS: An indicator variable taking value 1 if an individual is hospitalized during the period, and 0 otherwise.
4. NUMHOS: Number of hospital stays during the period.
5. NUMHOSDAY: Number of hospital days during the period.
6. ICU: An indicator variable taking value 1 if an individual receives care in intensive care unit (ICU) during the period, and 0 otherwise.
7. NUMICU: Number ICU episodes during the period.
8. NUMICUDAY: Number of ICU days during the period.

##### Chronic Disease Diagnosis

9. CHRDIAG: An indicator variable taking value 1 for any new chronic disease diagnosis (listed in Supplement Table S1), and 0 otherwise.

Using ICD-10 disease classification codes in the UPC database, we identified new diagnoses of major chronic diseases. For this, we use the diseases described by the Charlson comorbidities index (Sundararajan et al. 2004) and to this index we add diagnoses of hypertension to create our measure. Supplement Table S1 (below) shows the specific ICD-10 codes that we classify as reflecting the presence of a major chronic disease.

### 1.5. Statistical analysis

We use a regression discontinuity design (RDD), a quasi-experimental study design capable of yielding an unbiased local average treatment effect (LATE) in the absence of treatment randomization. In the specific context of our study, the copayment tier (and corresponding copayment amount) is the ‘treatment’ of interest, and assignment shifts discontinuously at two specific thresholds in the underlying continuous monthly earnings distribution (measured in units of MMWs): 2 and 5 MMWs. Because this deterministic assignment rule generates a discontinuity in the probability of exposure among individuals with essentially identical earnings on either side of the threshold (identical in the limit as one approaches the threshold), treatment assignment is ‘as-good-as-random’ for individuals in the neighborhood of the threshold, enabling causal inference (Lee and Lemieux 2010; Bor et al. 2014). Importantly, this framework assumes continuity on the conditional expectation functions for each of the potential outcomes at the threshold, this assumption implies no-manipulation of running variable and continuity of baseline variables at the threshold (Lee and Lemieux 2010; Calonico, Cattaneo, and Titiunik 2014; Imbens and Lemieux 2008), which we investigate and for which we show support in Section 2.2.

Following Moscoe et al. (*39*) and the potential outcome framework, the average causal effect (ACE) in the sharp RD (SRD) design is defined as:

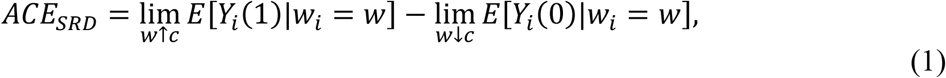

Where *Y*_*i*_(1) is an outcome of interest (outpatient use, inpatient use, 7-year survival, etc.) for individual *i*, when “exposed” (i.e. the worker with earnings just to the right of a threshold); *Y*_*i*_(*0*) is the outcome for individual *i* when “unexposed” (i.e. the workers with earnings just to the left of a threshold); and *w* is the continuous running variable (i.e. earnings in minimum wage units) in which there is a threshold or cut-off point *c* (i.e. 2 or 5) at which the probability of being exposed to a higher copayment changes discontinuously.

Our precise estimation strategies are as follows:

#### 1.5.1. RDD Estimation for ‘Contemporaneous’ Outpatient Service Use

We begin by using a RDD framework to estimate the contemporaneous causal relationship between copayment and outpatient service use in a given month. Specifically, for each threshold *c* (2 or 5), we estimate (1) using a standard local linear regression (Fan 2018). In particular, the estimate of *ACE*_*SRD*_ is given by:

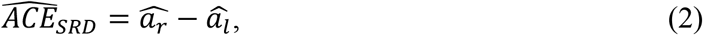

where

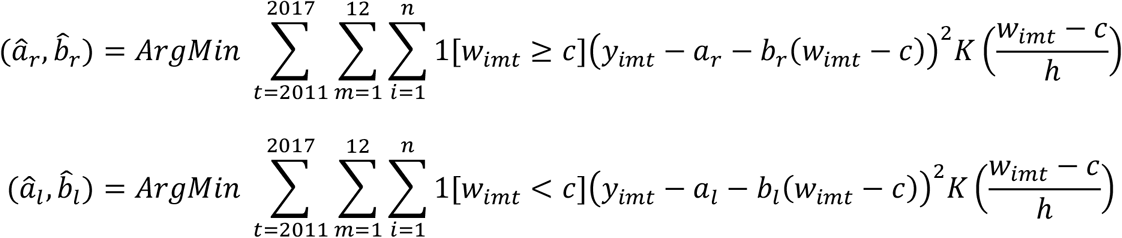

where *y*_*imt*_ is an outcome (outpatient services as well as each component of outpatient care described in Section 1.4: medical consultations, drugs, laboratory procedures, and diagnostic imaging procedures) for individual *i* in month *m* and year *t, h* is the robust bias-corrected ‘optimal’ sample bandwidths bandwidth (Calonico, Cattaneo, and Titiunik 2014; Calonico, Cattaneo, and Farrell 2020), and K(.) is the triangular Kernel. This estimation procedure restricts the sample to a distance *h* from either side of a threshold: *c-h* ≤ *w*_*imt*_ ≤ c *+ h*. The standard errors are adjusted for heteroskedasticity and clustering at the individual level.

#### 1.5.2. RDD Estimation for Chronic Disease Diagnosis

Next, we extend our statistical framework to study how these ‘contemporaneous’ changes in outpatient service use accumulate over time to influence the subsequent detection and diagnosis of chronic diseases. Specifically, we use ordinary least square (OLS) to estimate, for each threshold:

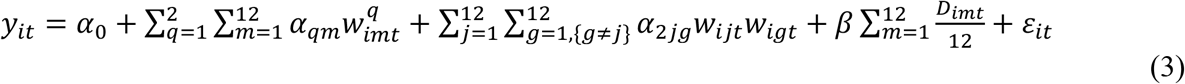

where *y*_*it*_ takes value 1 if the individual has been diagnosed of a new chronic disease within the year *t* or the 7 year period of the study, and 0 otherwise; *w*_*imt*_ is the running variable, the individual’s *i* earnings in month *m* and year *t*; *D*_*imt*_ is an indicator variable taking a value of 1 if *w*_*imt*_ is equal to or above above a threshold (2 or 5) and 0 otherwise – and 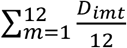 is the share of months in a year that an individual falls above the corresponding threshold; and *ε*_*imt*_ is an error term. The parameter *β* captures the effect of copayment on new chronic disease diagnoses. We restrict the sample to individuals within bandwidth *h* of the median wage 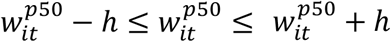, where 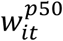 is the median of individual’s *i* monthly earnings in year *t*), and we again adjust standard errors for heteroskedasticity and clustering at the individual level. For this analysis, we only use individual-years with complete records for the 12 months of year *t*. Following Gelman and Imbens 2019, we include a second-order polynomial (given that higher order polynomials can lead to biased results).

#### 1.5.3. RDD Estimation for Inpatient Care or Emergency Room (ER) Use

Next, we study how the ‘contemporaneous’ changes in outpatient service use and chronic disease diagnosis accumulate over time to influence the use of potentially avoidable inpatient services. Specifically, we use ordinary least square (OLS) to estimate, for each threshold:

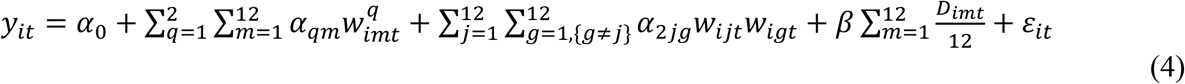

where *y*_*it*_ is an outcome related to inpatient or ER service use as (describe in Section 1.4) for individual *i* in year *t*; *w*_*imt*_ is the running variable, the individual’s *i* earnings in month *m* and year *t*; *D*_*imt*_ is an indicator variable taking a value of 1 if *w*_*imt*_ is equal to or above above a threshold (2 or 5) and 0 otherwise – and 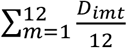 is the share of months in a year that an individual falls above the corresponding threshold; and *ε*_*it*_ is an error term. The parameter *β* captures the effect of copayment on new chronic disease diagnoses and inpatient care use. We restrict the sample to individuals within bandwidth *h* of the median wage 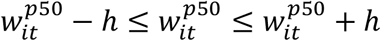 where 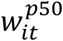 is the median of individual’s *i* monthly earnings in year *t*), and we again ajust standard errors for heteroskedasticity and clustering at the individual level. For this analysis, we only use individual-years with complete records for the 12 months of year *t*.

#### 1.5.4. Duration Analysis of Mortality Over Time

Following the approach of Bor et al. 2014, we study the survival of individuals facing different outpatient copayment rates over time by integrating our RDD into a duration analysis framework. This approach uses a semiparametric regression model to specify the mortality hazard (i.e., the instantaneous probability of death at time *t*, conditional on survival up to time *t*) as a function of the ‘running variable’ (in our case, monthly earnings) and time. Because monthly earnings vary over time, we use a time-dependent Cox model, which is appropriate with ‘external’ covariates (i.e., covariates vary as a function of time, independent of the failure time) (Cox 1972; Prentice and Kalbfleisch 1979; Cox 1975; Kalbfleisch and Prentice 2011). This is a plausible assumption if the RDD assumptions are otherwise met (as we examine, and for which we show support, in Section 2.2). Specifically, we estimate the causal hazard ratio (*CHR*), following Bor et al. 2014, as:

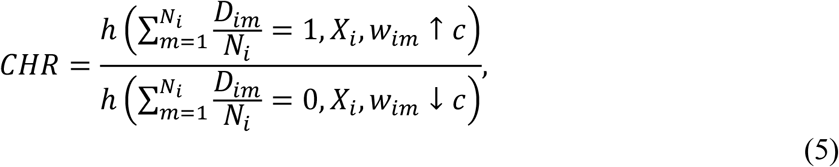

where:

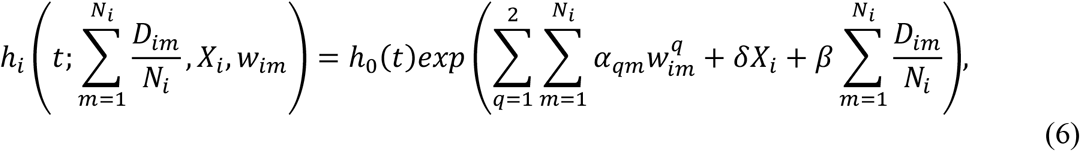

*h*_*i*_ (.) is the 7-year mortality hazard for individual *i*; *w*_*im*_is individual *i*’s earnings in month *m, m*∈ [1,84]; *D*_*im*_ is an indicator variable taking value 1 if *w*_*im*_ is equal to or greater than a threshold *c* (2 or 5 MMWs) and 0 otherwise, meaning that 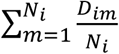 share of months in an N-month period that individual *i* falls above the c threshold; *N*_*i*_ is the number of months that individual *i* is observed in PILA database (i.e. number of months that individual *i* and his/her employer make payroll contributions); *X*_*i*_ is a vector of time-invariant individual characteristics (age in 2011, sex, enrollment with a public (vs. private) insurer, and geographic region of enrollment of which there are 5 in Colombia). We also include quadratic monthly earnings polynomials. The parameter *β* again captures the effect of higher copayments on the 7-year mortality hazard. We restrict the sample to individuals within bandwidth *h* of the median wage 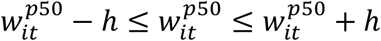, where 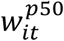 is the median of individual’s *i* monthly earnings over the period of observation). Follow Austin 2010, we estimate the absolute difference in 7-year mortality risk for both copayment thresholds using the time-dependent Cox model.

Finally, in addition to time-dependent Cox models, we also use several parametric survival models to estimate the causal hazard ratio of facing a higher-tier copayment. Specifically, we use Weibull and Exponential functions to identify the causal hazard ratio described in Equation (5).

#### 1.5.5. Elasticities

To facilitate both the interpretation of our results and comparisons of effect sizes between the two copayment thresholds (at 2 and 5 MMWs), we also estimate arc elasticities for each of the models previously described. For the analysis described in 1.5.1, we use the following:

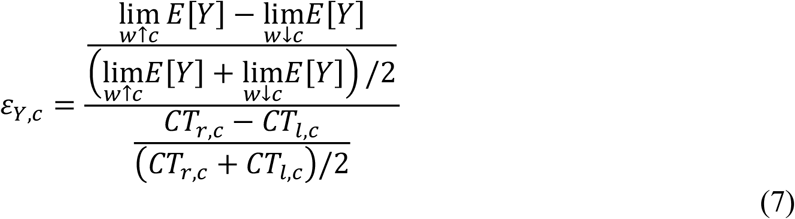

where *ε_Y,c_* is the arc elasticity of an outcome *Y* at threshold 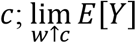 is the limit of the expected value of *Y Y,c* as earnings *w* approaches the threshold from above (in the earnings distribution); 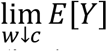 is the limit of the expected value of *Y* as earnings approaches the threshold from below (in the earnings distribution); *CT*_*r,c*_is the copayment value (in daily minimum wages) above the threshold (CT3 if c =5 or CT2 if c =2) and *CT*_*l,c*_is the copayment value (in daily minimum wages) below the threshold (CT2 if c =5 or CT1 if c =2).

For the analysis described in 1.5.2, 1.5.3, and 1.5.4, we use:

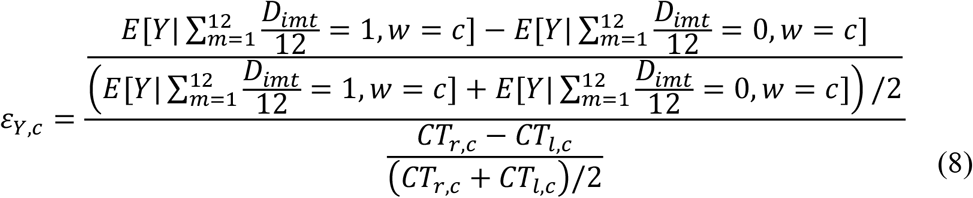

where 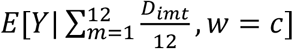 refer to the predictions of the regression or survival models indicated in (3), (4), and (5), and *CT*_*r,c*_ is defined as in (7).

### 2. Supplementary Text

#### 2.1. Descriptive Statistics

Supplement Table S2 shows summary statistics of Contributory Regime enrollees in our sample, both overall and by copayment tier. Among 12,850,726 individuals meeting inclusion criteria (i.e. all individuals enrolled in Contributory Regime for at least one month between January 2011 and December 2017, excluding individuals who reached the legal retirement age (57 for women and 62 for men) by 2011), there were 11,764,672 for at least one month in copayment tier 1 (below 2 MMWs), 4,128,676 for at least one month in tier 2 (between 2 and 5 MMW), and 1,406,560 for at least one month in tier 3 (above 5 MMWs).

#### 2.2. Evaluation of RDD Assumptions

##### 2.2.1. Tests for Covariate Continuity/Balance

Considering that the baseline characteristics are time invariant, but the running variable (i.e. monthly earnings) varies over time, we used randomization inference with false thresholds to test for true discontinuities in the distribution of the baseline characteristics at the threshold.

Supplement Figures S2, S3 and S4 then also show tests of continuity/balance across each of the thresholds using individual characteristics that cannot be influenced by cost-sharing: age, sex, enrollment with a public insurer (which can only change annually at the time of ‘open enrollment’), and region of enrollment. Supplement Figures S2 and S3 show balance, or the absence of a discontinuity, in demographic characteristics across both cost-sharing thresholds. Then, to test for statistically significant differences across each threshold, we randomly draw 500 false thresholds and re-estimate the same local linear regressions (using the demographic characteristics as dependent variables). Supplement Figure S4 shows that the null hypothesis of continuity in these variables across each threshold is not rejected – a result consistent with the assumptions of the RDD model.

##### 2.2.2. McCrary Density Test for Sorting Around the Thresholds

Our RDD estimation (and the estimation frameworks which build on them – described in detail in Section 1) assumes no manipulation of the ‘running variable’ (in our case, that individuals do not manipulate the reporting of their earnings in the PILA system to gain lower cost-sharing requirements). Such manipulation would be evident as a mass-point of individuals just below a threshold (2 or 5 in the distribution of *w*_*im*_), which we formally test using the McCrary density test (McCrary 2008). Supplement Figure S5 shows the density of observations across the earnings distribution. We then implement a McCrary 2008 density test using randomization inference at each copayment threshold, finding no statistically significant evidence of a discontinuity in sample density at either – a result again consistent with the assumptions of the RDD model.

#### 2.3. Cost-Sharing and Monthly Outpatient Care Service Use

Figure 1 in the paper shows total outpatient care service use across both copayment thresholds in the distribution of earnings. Breaking outpatient services into its components (medical consultations, drugs, laboratory procedures, and diagnostic imaging procedures), Supplement Figures S6 and S7, and Supplement Tables S3 and S4 show equivalent analyses for each component at each threshold (2 MMW and 5 MMW).

#### 2.4. Effect of copayment on Annual Outpatient Care Services

For comparability with subsequent analysis, we also estimate the effect of copayments on annual outpatient care services using Equation (3) where *yit* represents annual measures of total outpatient services and the number of each of its components (medical consultations, drugs, laboratory procedures, and diagnostic imaging procedures). Figure 2 and Supplement Tables S5 and S6 show these results. In all cases (and at both the 2 MMW and 5 MMW copayment threshold), the estimated copayment effects are negative and statistically significant. To aid in interpretation, the tables also report arc elasticities computed using (8).

#### 2.5. Outpatient Cost Sharing and Subsequent Diagnoses of Chronic Disease and Hospital Care ‘Offsets’

Extending our RDD estimation framework to examine how differences in outpatient cost-sharing (and service use) accumulate over time, contributing to the detection of chronic disease, Supplement Tables S7 and S8 report estimates from Equation (3) which correspond to the results shown in paper Figure 3A. Separate tables show results for each cost-sharing threshold (2 MMWs and 5 MMWs). In general, over both shorter and longer periods of time (1 and 7 years), they consistently show a relationship between greater outpatient cost-sharing and lower probabilities of detecting chronic diseases. To aid in interpretation, we also report arc elasticities computed using (8).

Similarly, we use the same estimation framework (Regression (4)) to estimate the effect of greater outpatient cost-sharing (and less outpatient service use and subsequent chronic disease diagnosis) on the use of hospital and emergency care. Specifically, Supplement Tables S9-S12 below show estimates that correspond to the results in paper Figure 3B (and extend them on the intensive margin – that is, the number of services used, not just probability of use). In general, across cost-sharing thresholds (at 2 and 5 MMWs), and sample bandwidths (0.5 and 0.3 MMWs), there is consistently a relationship between greater outpatient care cost-sharing and more emergency room visits, more (and longer) inpatient stays, and greater (and longer) use of intensive care units. These results suggest the presence of important hospital care “offsets” – that is, the use of services that are potentially avoidable through appropriate use of outpatient care. Again, to aid in interpretation, we also report arc elasticities computed using (8).

#### 2.6. Outpatient Cost-Sharing and Subsequent Mortality/Survival

Finally, Table S13 show causal hazard ratio (CHR) estimates obtained using extended Cox hazard models as shown in (5) and (6). These estimates underly the results shown in Figures 4A and 4B, which we also present in Tables S14 and S15 by analyzing sensitivity to different hazard models (parametric models with Exponential and Weibull distributions for the survivor function). We observe that the mortality hazard ratios increase with the amount of time that individuals are subject to copayment requirements in the formal sector. Again, to aid in interpretation, we also report arc elasticities computed using (8).

**Fig. S1.**
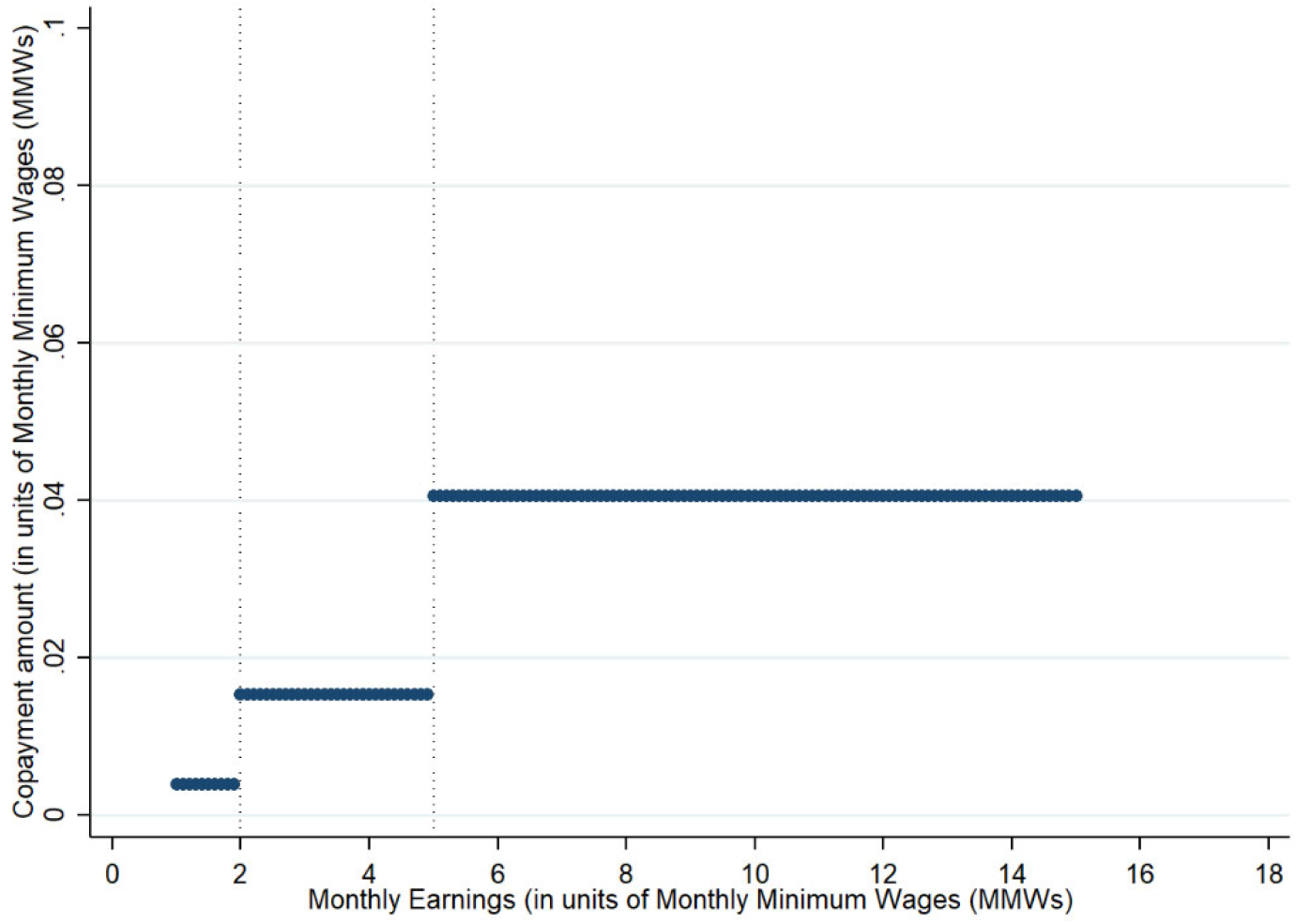
Outpatient Service Copayment Requirements for Formal-Sector Employees in Colombia. Contributory Regime enrollees face a step-function copayment for outpatient services that varies with earnings (measured in Monthly Minimum Wages (MMWs)). There are three copayment tiers (CT); CT1: 11.7% of a daily minimum wage, which roughly corresponds to 3,400 Colombian Pesos (COP $) (or 0.92 United States Dollars (USD $)) in 2020; CT2: 46.1% of a daily minimum wage, which roughly corresponds to COP $ 13,500 (USD $ 3.65) in 2020; and CT3: 121.5% of a daily minimum wage, which roughly corresponds to COP $ 35,600 (USD $ 9.62) in 2020.

**Fig. S2.**
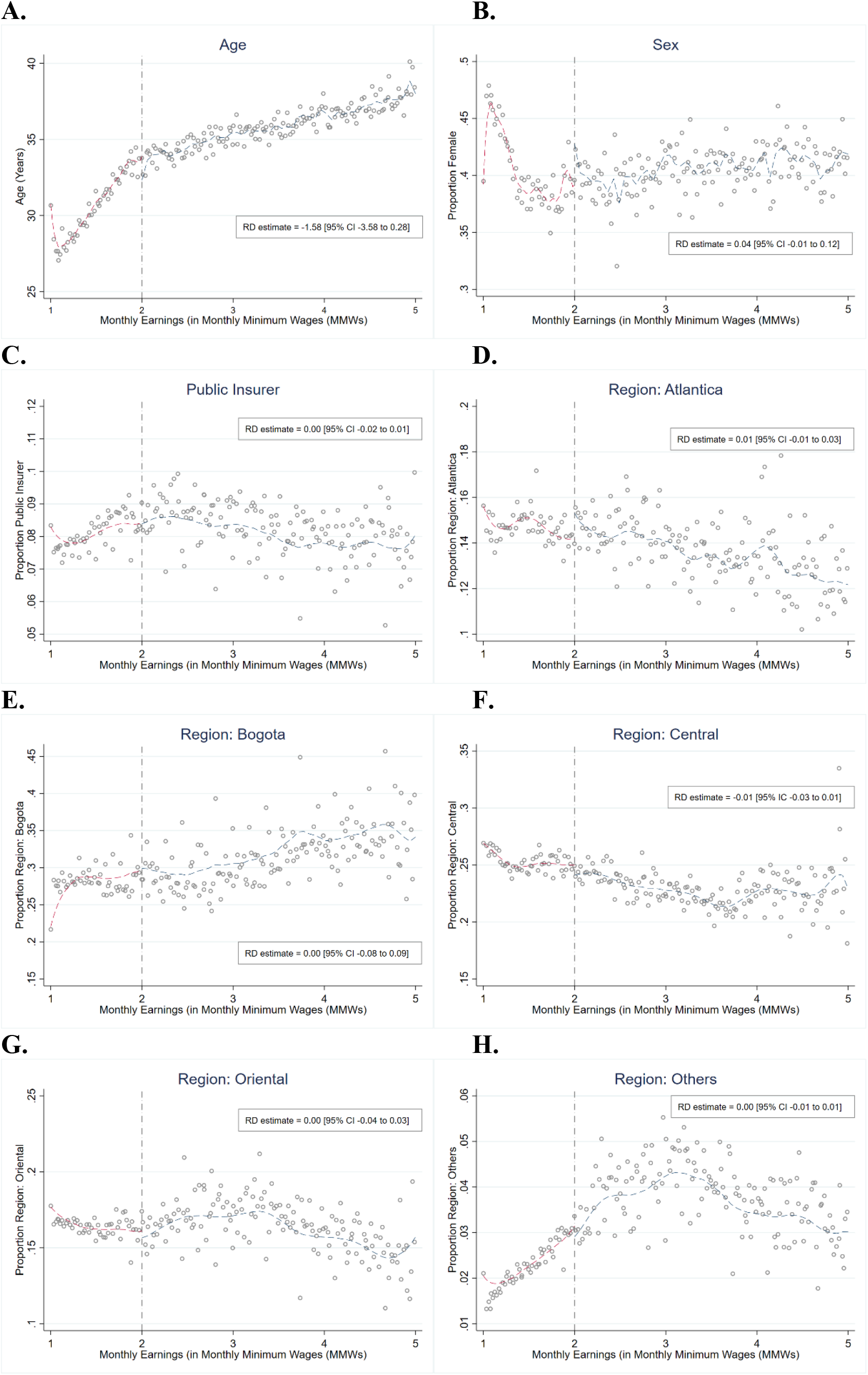
Balance in Characteristics across the Copayment Threshold (at 2 Monthly Minimum Wages). Regression discontinuity (RD) graphs of balance, or the absence of a discontinuity, in demographic characteristics with local linear smoothing for both sides of the threshold at 2 monthly minimum wages. Each panel shows a demographic characteristic: A. Age in years; B. Proportion of women; C, Proportion of Public Insurer; D. Proportion in Region: Atlántica; E. Proportion in Region: Bogotá; F. Proportion in Region: Central; G. Proportion in Region: Oriental; and H. Proportion in Region: Others. We randomly draw 500 false thresholds and estimate the local linear regressions using the demographic characteristics as dependent variable to test for statistical significance.

**Fig. S3.**
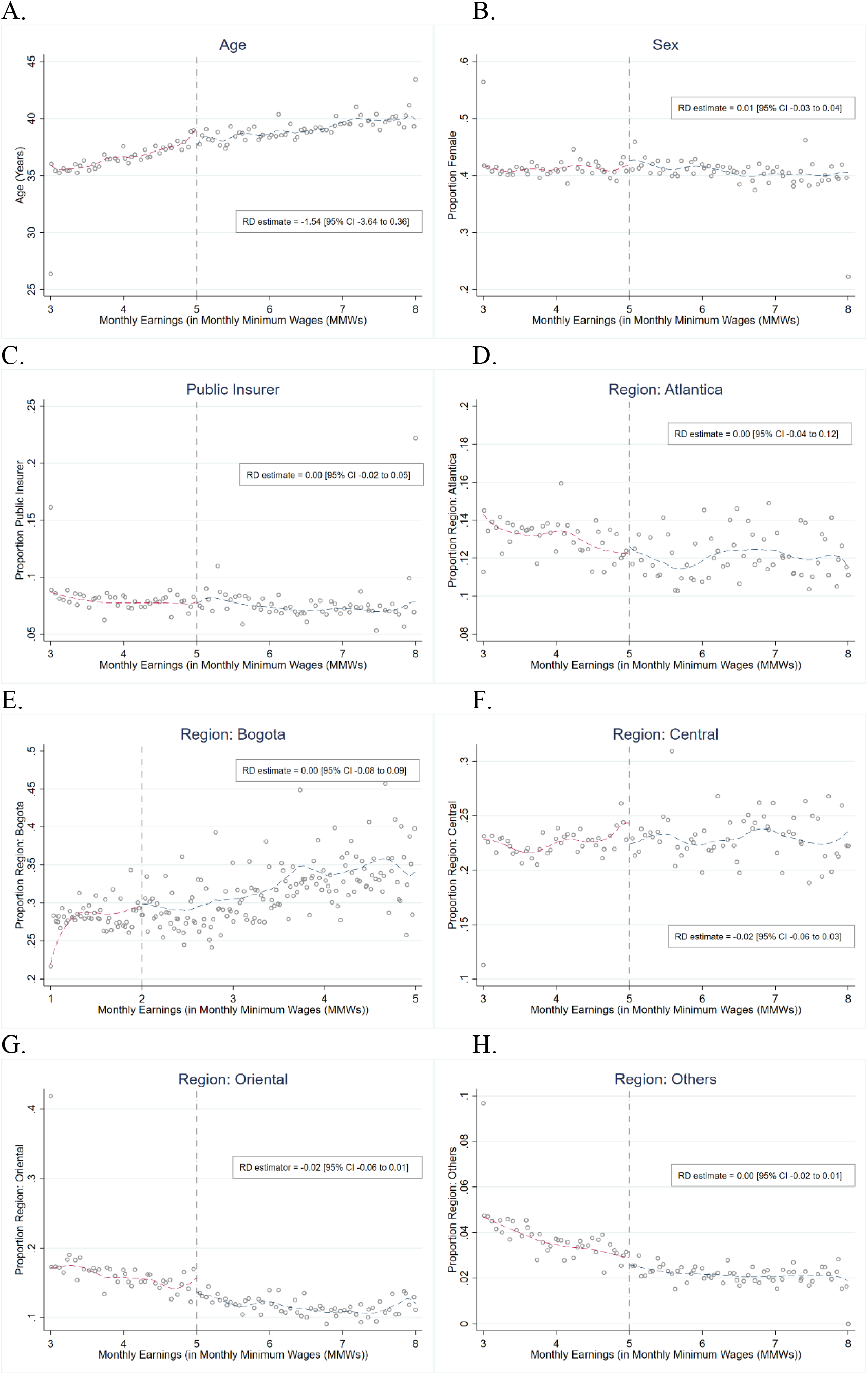
Balance in Characteristics across the Copayment Threshold (at 5 Monthly Minimum Wages). Regression discontinuity (RD) graphs of balance, or the absence of a discontinuity, in demographic characteristics with local linear smoothing for both sides of the threshold at 5 monthly minimum wages. Each panel shows a demographic characteristic: A. Age in years; B. Proportion of women; C, Proportion of Public Insurer; D. Proportion in Region: Atlántica; E. Proportion in Region: Bogotá; F. Proportion in Region: Central; G. Proportion in Region: Oriental; and H. Proportion in Region: Others. We randomly draw 500 false thresholds and estimate the local linear regressions using the demographic characteristics as dependent variable to test for statistical significance.

**Fig. S4.**
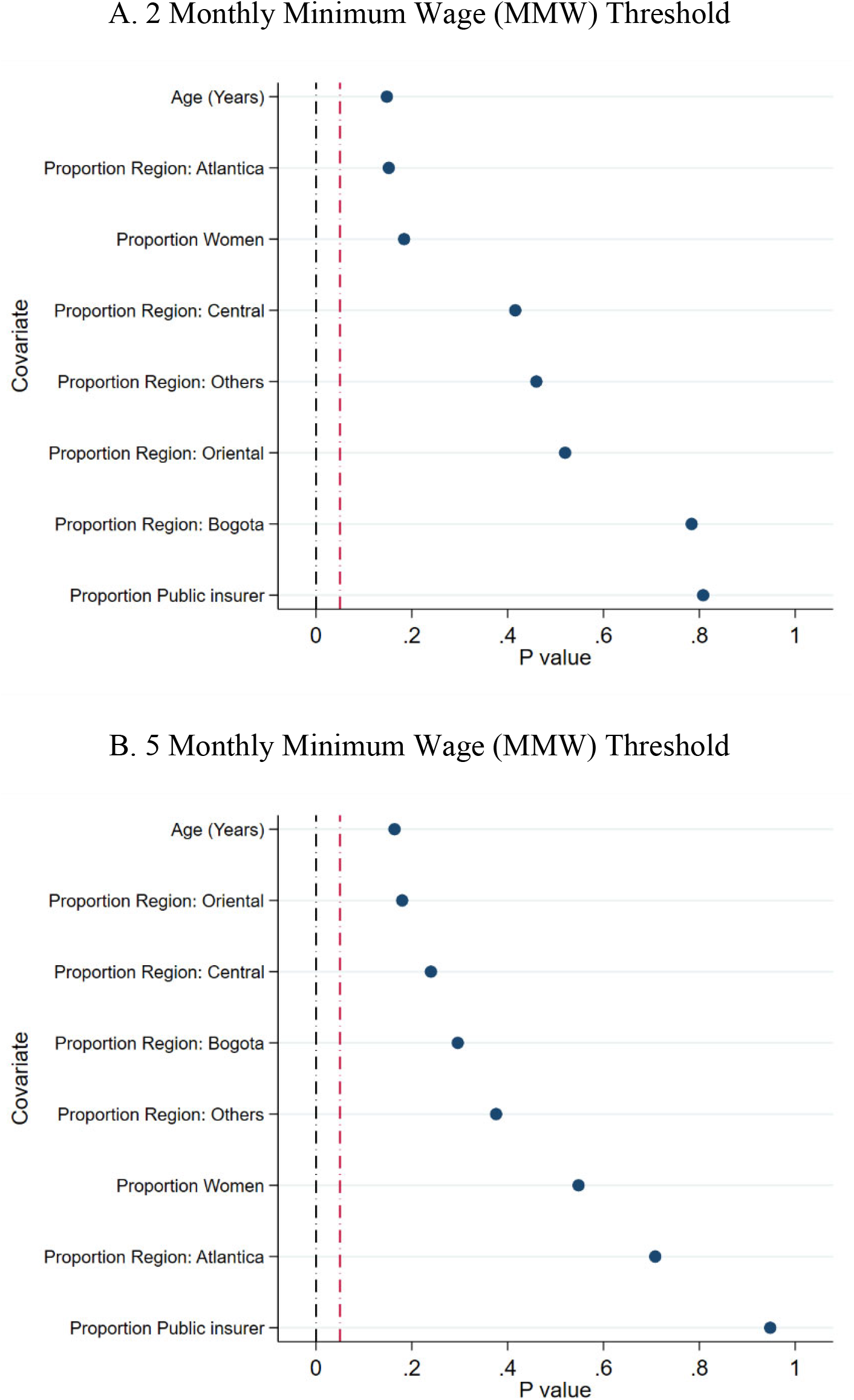
Covariate Falsification Significance Tests. Tests of continuity of baseline covariates at the 2 Monthly Minimum Wage (MMW) Threshold (**A**) and the 5 Monthly Minimum Wage (MMW) Threshold (**B**). We randomly draw 500 false thresholds and estimate the local linear regressions using the demographic characteristics as dependent variable to test for statistical significance.

**Fig. S5.**
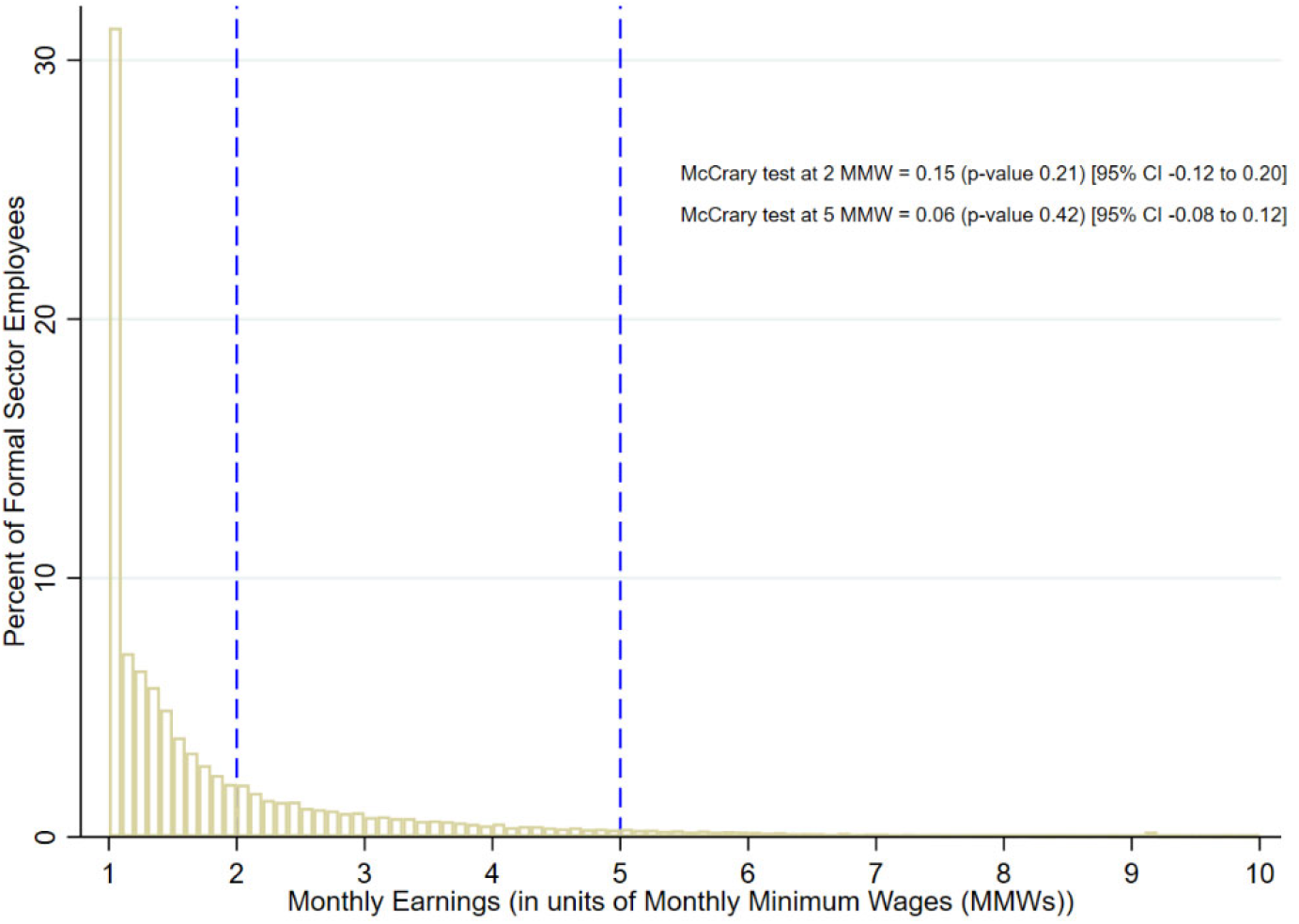
McCrary (2008) Density Test for Running Variable Manipulation. Histogram of the distribution of observations across the monthly earning (in unit of Monthly Minimum Wages) distribution. We implement the McCrary density test using randomization inference with 500 false thresholds at each copayment threshold.

**Fig. S6.**
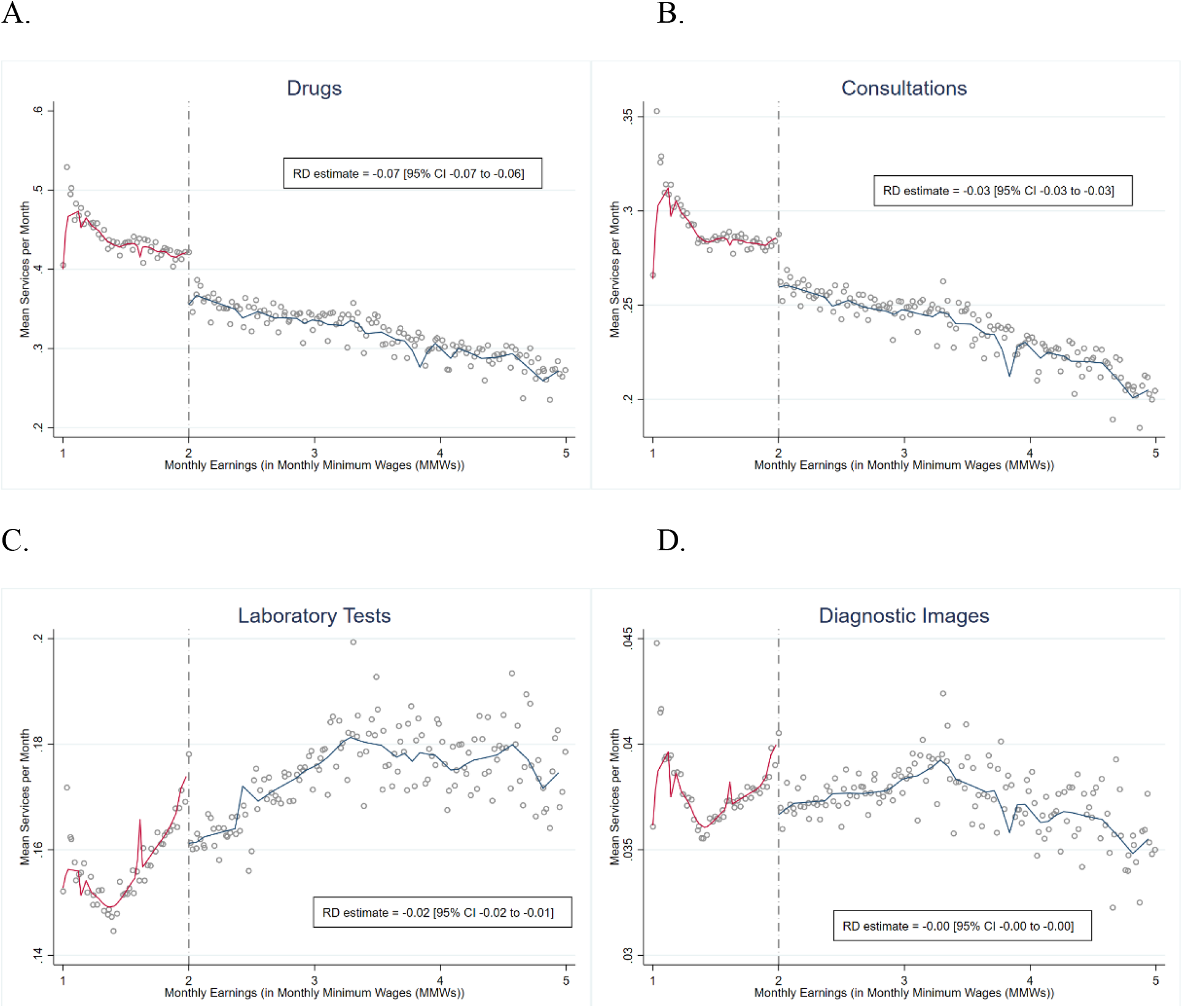
Outpatient Service Use by Type across the Copayment Threshold (at 2 Monthly Minimum Wages (MMWs)). Regression discontinuity (RD) graphs of outpatient service use by type with local linear smoothing for both sides of the threshold at 2 monthly minimum wages. (A) Drugs. (B) Consultations. (C) Laboratory Tests. (D) Diagnostic Images. Vertical axis show means of services used per month for all formal workers between 2011 and 2017. We use local linear regression with robust bias-corrected ‘optimal’ sample bandwidths, standard errors adjusting for heteroskedasticity and clustering at the individual level, to estimate the effect of cost-sharing on the total outpatient service use.

**Fig. S7.**
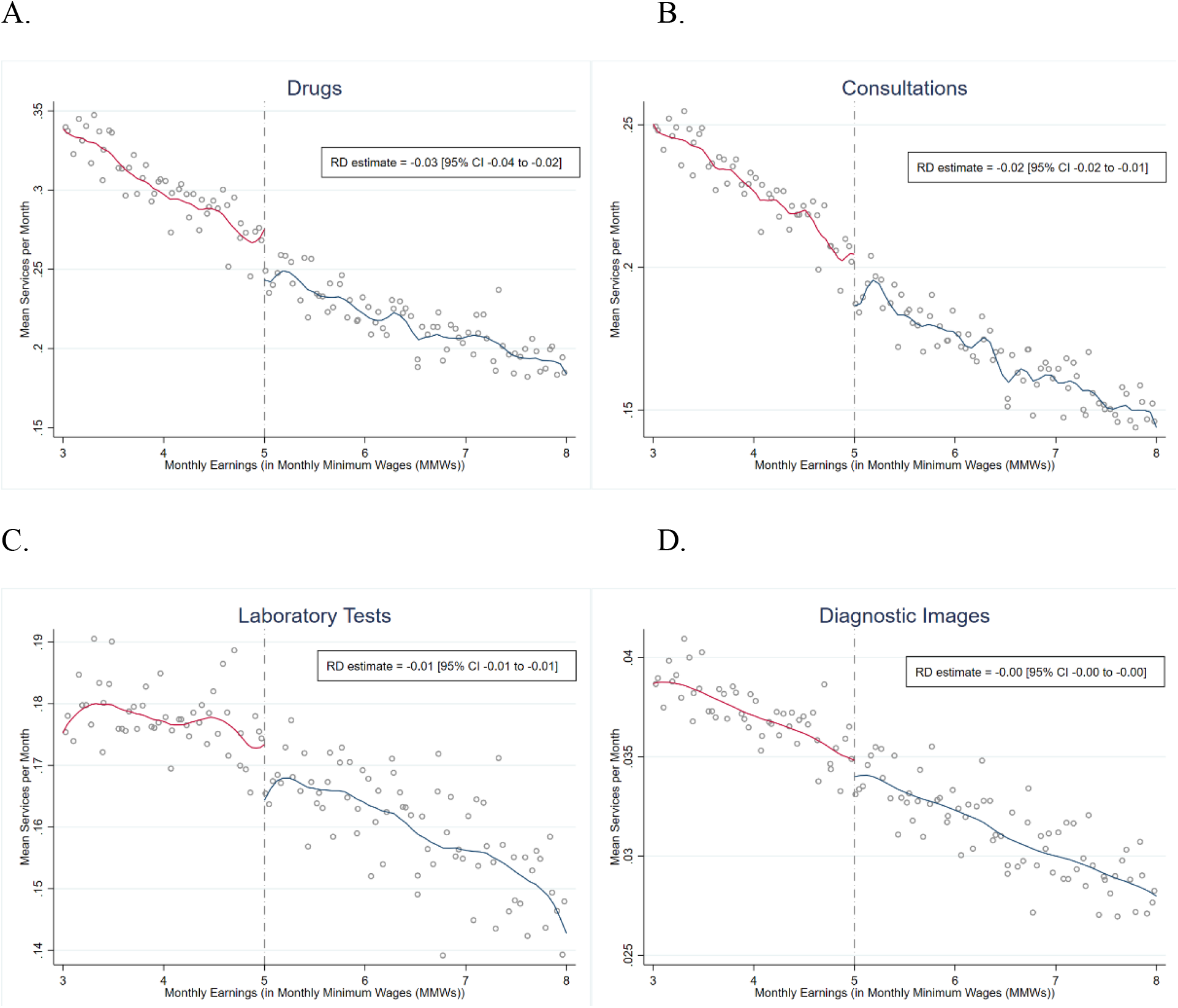
Outpatient Service Use by Type across the Copayment Threshold (at 5 Monthly Minimum Wages (MMWs)) Regression discontinuity (RD) graphs of outpatient service use by type with local linear smoothing for both sides of the threshold at 5 monthly minimum wages. (A) Drugs. (B) Consultations. (C) Laboratory Tests. (D) Diagnostic Images. Vertical axis show means of services used per month for all formal workers between 2011 and 2017. We use local linear regression with robust bias-corrected ‘optimal’ sample bandwidths, standard errors adjusting for heteroskedasticity and clustering at the individual level, to estimate the effect of cost-sharing on the total outpatient service use.

**Table S1.** ICD-10 Codes Used to Identify Chronic Disease Diagnosis in *Base del Estudio de Suficiencia de la Unidad Por Capitación*, or UPC Database. Arterial hypertension codes are not included in the Charlson comorbidities index. However, we include them due to the high prevalence of patients with hypertension. ICD-10: International Classification of Diseases, 10th Revision.

| Disease | ICD-10 Codes |
| --- | --- |
| Acute myocardial infarction | I21, I22, I252 |
| Congestive heart failure | I50 |
| Peripheral vascular disease | I71, I790, I739, R02, Z958, Z959 |
| Cerebral vascular accident | I60, I61, I62, I63, I65, I66, G450, G451, G452, G458, G459, G46, I64, G454, I670, I671, I672, I674, I675, I676, I677, I678, I679, I681, I682, I688, I69 |
| Dementia | F00, F01, F02, F051 |
| Pulmonary disease | J40, J41, J42, J44, J43, J45, J46, J47, J67, J44, J60, J61, J62, J63, J66, J64, J65 |
| Connective tissue disorder | M32, M34, M332, M053, M058, M059, M060, M063, M069, M050, M052, M051, M353 |
| Peptic ulcer | K25, K26, K27, K28 |
| Liver disease | K702, K703, K73, K717, K740, K742, K746, K743, K744, K745 |
| Diabetes | E109, E119, E139, E149, E101, E111, E131, E141, E105, E115, E135, E145 |
| Diabetes complications | E102, E112, E132, E142 E103, E113, E133, E143 E104, E114, E134, E144 |
| Paraplegia | G81, G041, G820, G821, G822 |
| Renal disease | N03, N052, N053, N054, N055, N056, N072, N073, N074, N01, N18, N19, N25 |
| Cancer | C0, C1, C2, C3, C40, C41, C43, C45, C46, C47, C48, C49, C5, C6, C70, C71, C72, C73, C74, C75, C76, C80, C81, C82, C83, C84, C85, C883, C887, C889, C900, C901, C91, C92, C93, C940, C941, C942, C943, C9451, C947, C95, C96 |
| Metastatic cancer | C77, C78, C79, C80 |
| Severe liver disease | K729, K766, K767, K721 |
| Human Immunodeficiency Virus | B20, B21, B22, B23, B24 |
| Arterial hypertension | I10, I11, I12, I13, I14, I15 |

**Table S2.** Descriptive Statistics. Descriptive statistics of main variables for all individuals enrolled in Contributory Regime for at least one month between January 2011 and December 2017, excluding individuals who reached the legal retirement age (57 for women and 62 for men) by 2011. Standard errors in parentheses. OUT: annual average of all outpatient services used; DRUGS: annual average of drugs used; CONS: annual average of medical consultations used; LAB: annual average of laboratory procedures used; IMAG: annual average of diagnostic imaging procedures used; ER: is an indicator variable taking value 1 if the worker visits the Emergency Room (ER) and 0 otherwise in a year; NUMER: annual average of visits to ER; HOS: is an indicator variable taking value 1 if the worker is hospitalized in a year and 0 otherwise; NUMHOS: annual average of hospitalizations; NUMHOSDAY: annual average of days of hospital stay; ICU: is an indicator variable taking value 1 if the worker is hospitalized in intensive care unit (ICU) in a year and 0 otherwise; NUMICU: annual average of hospitalizations in ICU; NUMICUDAY: annual average of days of hospital stay in ICU.

| VARIABLES | (1)<br>Full sample | (2)<br>$1 \leq w_{imt} < 2$ | (3)<br>$2 \leq w_{imt} \leq 5$ | (4)<br>$w_{imt} > 5$ |
| --- | --- | --- | --- | --- |
| Age (in Years) | 31.1<br>(0.00316) | 30.49<br>(0.00326) | 32.69<br>(0.00518) | 36.22<br>(0.00868) |
| Share Women (%) | 40.79<br>(0.0137) | 40.55<br>(0.0143) | 38.16<br>(0.0239) | 38.8<br>(0.0411) |
| Share Enrolled in the Public Plan (%) | 8.079<br>(0.0076) | 8.11<br>(0.00796) | 7.596<br>(0.013) | 6.789<br>(0.0212) |
| Share by Geographical Region (%) |  |  |  |  |
| Atlántica | 14.89<br>(0.00993) | 15.08<br>(0.0104) | 13.82<br>(0.017) | 12.83<br>(0.0282) |
| Bogotá | 26.38<br>(0.0123) | 25.18<br>(0.0127) | 31.15<br>(0.0228) | 39.01<br>(0.0411) |
| Central | 25.7<br>(0.0122) | 26.01<br>(0.0128) | 24.11<br>(0.0211) | 21.58<br>(0.0347) |
| Oriental | 16.79<br>(0.0104) | 17.22<br>(0.011) | 16.19<br>(0.0181) | 13.73<br>(0.029) |
| Pacífica | 14.01<br>(0.00968) | 14.29<br>(0.0102) | 11.87<br>(0.0159) | 10.39<br>(0.0257) |
| Others | 2.22<br>(0.00411) | 2.216<br>(0.00429) | 2.858<br>(0.0082) | 2.468<br>(0.0131) |
| 7-year Cumulative Mortality Risk (%) | 0.658<br>(0.00226) | 0.658<br>(0.00236) | 0.561<br>(0.00368) | 0.548<br>(0.00622) |
| Chronic Disease Diagnosis (%) | 16.47<br>(0.0103) | 16.08<br>(0.0107) | 21.27<br>(0.0201) | 21.37<br>(0.0346) |
| Earnings (in Monthly Minimum Wages (MMWs)) | 1.831<br>(0.00054) | 1.435<br>(0.00024) | 2.708<br>(0.00083) | 5.753<br>(0.00323) |
| Annual Outpatient Services Use |  |  |  |  |
| OUT (mean) | 7.023<br>(0.00321) | 7.037<br>(0.00333) | 7.869<br>(0.00558) | 6.772<br>(0.00938) |
| DRUGS (mean) | 3.205<br>(0.00177) | 3.252<br>(0.00185) | 3.451<br>(0.00307) | 2.718<br>(0.00511) |
| CONS (mean) | 2.215<br>(0.000904) | 2.233<br>(0.000949) | 2.488<br>(0.00153) | 2.055<br>(0.00244) |
| LAB (mean) | 1.305<br>(0.000924) | 1.26<br>(0.000939) | 1.576<br>(0.00168) | 1.66<br>(0.00307) |
| IMAG (mean) | 0.298<br>(0.000215) | 0.292<br>(0.000222) | 0.355<br>(0.000376) | 0.339<br>(0.000592) |
| Annual Inpatient Services Use |  |  |  |  |
| ER (%) | 46.05<br>(0.0139) | 46.21<br>(0.0145) | 58.23<br>(0.0243) | 52.65<br>(0.0421) |
| NUMER (mean) | 0.423<br>(0.000247) | 0.435<br>(0.000264) | 0.454<br>(0.0004) | 0.343<br>(0.000563) |
| HOS (%) | 12.73<br>(0.0093) | 12.74<br>(0.00972) | 16.72<br>(0.0184) | 14.98<br>(0.0301) |
| NUMHOS (mean) | 0.195 | 0.2 | 0.189 | 0.156 |
|  | (0.00059) | (0.000626) | (0.000857) | (0.00134) |
| NUMHOSDAY (mean) | 0.0737 | 0.075 | 0.0759 | 0.0624 |
|  | (0.0000972) | (0.000103) | (0.000146) | (0.000223) |
| ICU (%) | 1.074 | 1.089 | 1.268 | 1.072 |
|  | (0.00287) | (0.00303) | (0.00551) | (0.00868) |
| NUMICU (mean) | 0.0311 | 0.0319 | 0.0274 | 0.0221 |
|  | (0.000207) | (0.000217) | (0.000314) | (0.000492) |
| NUMICUDAY (mean) | 0.00487 | 0.00499 | 0.00434 | 0.00351 |
|  | (0.0000235) | (0.0000244) | (0.0000338) | (0.0000468) |
| Observations | 12,850,726 | 11,764,672 | 4,128,676 | 1,406,560 |

**Table S3.** The Effect of Cost-Sharing on Outpatient Service Use, Total and by Type (at the 2 Monthly Minimum Wage (MMW) Threshold). Local linear regressions (LLR) estimates obtained using Equation (2) (dependent variables are the number of services and procedures, total and by type, shown at the top of each column). We use the full sample of individuals enrolled in the Contributory Regime for at least one month between January 2011 and December 2017, excluding individuals who reached the legal retirement age (57 for women and 62 for men) by 2011, to estimate the effect of cost-sharing at the 2 Monthly Minimum Wage (MMW) threshold on the outpatient service use. We use robust bias-corrected ‘optimal’ sample bandwidths and adjust standard errors for heteroskedasticity and clustering at the individual level.

| Outpatient<br>Services Use | (1)<br>Total<br>Outpatient | (2)<br>Drugs | (3)<br>Consultations | (4)<br>Diagnostic<br>Images | (5)<br>Laboratory<br>Tests |
| --- | --- | --- | --- | --- | --- |
| Outpatient<br>Cost-Sharing Effect | -0.115***<br>(0.00491) | -0.0667***<br>(0.00285) | -0.0277***<br>(0.00127) | -0.00399***<br>(0.000298) | -0.0157***<br>(0.00124) |
| Observations | 14,410,541 | 13,633,739 | 16,188,180 | 21,437,670 | 25,676,933 |
| Optimal Bandwidth | 0.08 | 0.08 | 0.09 | 0.12 | 0.15 |
| Mean below threshold | 0.93 | 0.42 | 0.29 | 0.04 | 0.18 |
| Mean above threshold | 0.81 | 0.36 | 0.26 | 0.04 | 0.16 |
| Elasticity | -0.11 | -0.14 | -0.09 | -0.09 | -0.08 |

\*\*\* p<0.01, \*\* p<0.05, \* p<0.1

**Table S4.** The Effect of Cost-Sharing on Outpatient Service Use, Total and by Type (at the 5 Monthly Minimum Wage (MMW) Threshold). Local linear regressions (LLR) estimates obtained using Equation (2) (dependent variables are the number of services and procedures, total and by type, shown at the top of each column). We use the full sample of individuals enrolled in the Contributory Regime for at least one month between January 2011 and December 2017, excluding individuals who reached the legal retirement age (57 for women and 62 for men) by 2011, to estimate the effect of cost-sharing at the 5 Monthly Minimum Wage (MMW) threshold on the outpatient service use. We use robust bias-corrected ‘optimal’ sample bandwidths and adjust standard errors for heteroskedasticity and clustering at the individual level.

| Outpatient Services Use | (1)<br>Total Outpatient | (2)<br>Drugs | (3)<br>Consultations | (4)<br>Diagnostic Images | (5)<br>Laboratory Tests |
| --- | --- | --- | --- | --- | --- |
| Outpatient Cost-Sharing Effect | -0.0624***<br>(0.00772) | -0.0323***<br>(0.00376) | -0.0181***<br>(0.00213) | -0.000871**<br>(0.000367) | -0.00894***<br>(0.00207) |
| Observations | 4,247,275 | 5,335,053 | 3,908,669 | 15,132,618 | 10,182,542 |
| Optimal Bandwidth | 0.16 | 0.21 | 0.15 | 0.59 | 0.40 |
| Mean below threshold | 0.69 | 0.28 | 0.20 | 0.03 | 0.17 |
| Mean above threshold | 0.63 | 0.24 | 0.19 | 0.03 | 0.16 |
| Elasticity | -0.10 | -0.14 | -0.10 | -0.03 | -0.06 |

\*\*\* p<0.01, \*\* p<0.05, \* p<0.1

**Table S5.** Cumulative Effect of Cost-Sharing on Outpatient Service Use, Total and by Type (at the 2 Monthly Minimum Wage (MMW) Threshold). Effect of greater outpatient cost-sharing on the annual use of total outpatient services and its components (number of drugs, consultations, laboratory tests, and diagnostic images) at the 2 Monthly Minimum Wage (MMW) Threshold. We use a sample of individuals with median monthly earnings within 0.5 (**A**) and 0.3 (**B**) MMWs of the threshold, and who worked during the entire year (12 months). We use ordinary least square with a second-order polynomial of the monthly earnings, and we compute heteroskedastic standard errors clustered at the individual level. We include a vector of time-invariant individual characteristics (age in 2011; sex; enrollment with a public (vs. private) insurer; and geographic region of enrollment, of which there are 5 in Colombia).

|  | (1) | (2) | (3) | (4) | (5) |
| --- | --- | --- | --- | --- | --- |
| Outpatient Services Use | Total Outpatient | Drugs | Consultations | Diagnostic Images | Laboratory Tests |
| A. 0.5 Monthly Minimum Wage (MMW) Bandwidth |  |  |  |  |  |
| Outpatient Cost-Sharing Effect | -1.371***<br>(0.0571) | -0.871***<br>(0.0332) | -0.349***<br>(0.0153) | -0.0235***<br>(0.00395) | -0.127***<br>(0.0172) |
| Observations | 5,220,007 | 5,220,007 | 5,220,007 | 5,220,007 | 5,220,007 |
| Mean below threshold | 11.50 | 5.340 | 3.559 | 0.486 | 2.111 |
| Mean above threshold | 10.13 | 4.469 | 3.209 | 0.463 | 1.985 |
| Elasticity | -0.107 | -0.149 | -0.0867 | -0.0417 | -0.0519 |
| B. 0.3 Monthly Minimum Wage (MMW) Bandwidth |  |  |  |  |  |
| Outpatient Cost-Sharing Effect | -1.332***<br>(0.0663) | -0.875***<br>(0.0382) | -0.333***<br>(0.0181) | -0.0167***<br>(0.00441) | -0.108***<br>(0.0203) |
| Observations | 2,996,596 | 2,996,596 | 2,996,596 | 2,996,596 | 2,996,596 |
| Mean below threshold | 11.48 | 5.313 | 3.547 | 0.487 | 2.133 |
| Mean above threshold | 10.15 | 4.439 | 3.214 | 0.471 | 2.025 |
| Elasticity | -0.103 | -0.151 | -0.0827 | -0.0293 | -0.0436 |

\*\*\* p<0.01, \*\* p<0.05, \* p<0.1

**Table S6.** Cumulative Effect of Cost-Sharing on Outpatient Service Use, Total and by Type (at the 5 Monthly Minimum Wage (MMW) Threshold). Effect of greater outpatient cost-sharing on the annual use of total outpatient services and its components (number of drugs, consultations, laboratory tests, and diagnostic images) at the 5 Monthly Minimum Wage (MMW) Threshold. We use a sample of individuals with median monthly earnings within 0.5 (**A**) and 0.3 (**B**) MMWs of the threshold, and who worked during the entire year (12 months). We use ordinary least square with a second-order polynomial of the monthly earnings, and we compute heteroskedastic standard errors clustered at the individual level. We include a vector of time-invariant individual characteristics (age in 2011; sex; enrollment with a public (vs. private) insurer; and geographic region of enrollment, of which there are 5 in Colombia).

|  | (1) | (2) | (3) | (4) | (5) |
| --- | --- | --- | --- | --- | --- |
| Outpatient Services Use | Total Outpatient | Drugs | Consultations | Diagnostic Images | Laboratory Tests |
| A. 0.5 Monthly Minimum Wage (MMW) Bandwidth |  |  |  |  |  |
| Outpatient Cost-Sharing Effect | -0.261***<br>(0.0874) | -0.231***<br>(0.0475) | -0.0495**<br>(0.0226) | 0.00489<br>(0.00576) | 0.0149<br>(0.0315) |
| Observations | 824,970 | 824,970 | 824,970 | 824,970 | 824,970 |
| Mean below threshold | 8.189 | 3.226 | 2.416 | 0.422 | 2.125 |
| Mean above threshold | 7.928 | 2.995 | 2.366 | 0.426 | 2.140 |
| Elasticity | -0.0360 | -0.0827 | -0.0230 | 0.0128 | 0.00774 |
| B. 0.3 Monthly Minimum Wage (MMW) Bandwidth |  |  |  |  |  |
| Outpatient Cost-Sharing Effect | -0.376***<br>(0.0989) | -0.281***<br>(0.0538) | -0.0856***<br>(0.0256) | -0.00319<br>(0.00656) | -0.00612<br>(0.0358) |
| Observations | 492,492 | 492,492 | 492,492 | 492,492 | 492,492 |
| Mean below threshold | 8.152 | 3.220 | 2.403 | 0.420 | 2.109 |
| Mean above threshold | 7.777 | 2.940 | 2.317 | 0.417 | 2.103 |
| Elasticity | -0.0524 | -0.101 | -0.0403 | -0.00847 | -0.00323 |

\*\*\* p<0.01, \*\* p<0.05, \* p<0.1

**Table S7.** Cumulative Effect of Outpatient Cost-Sharing on Chronic Disease Diagnosis (1-Year and 7-Year Horizons, at the 2 Monthly Minimum Wage (MMW) Threshold). Effect of greater outpatient cost-sharing on the probabilities (percentage points) of detecting chronic diseases for the same year and for all 7 years in the sample at the 2 Monthly Minimum Wage (MMW) Threshold. We use a sample of individuals with median monthly earnings within 0.5 and 0.3 MMWs of the threshold, and who worked during the entire year (12 months). We use ordinary least square with a second-order polynomial of the monthly earnings, and we compute heteroskedastic standard errors clustered at the individual level. We include a vector of time-invariant individual characteristics (age in 2011; sex; enrollment with a public (vs. private) insurer; and geographic region of enrollment, of which there are 5 in Colombia).

| Chronic Disease<br>Diagnosis | (1)<br>1 year | (2)<br>7 years | (3)<br>1 year | (4)<br>7 years |
| --- | --- | --- | --- | --- |
| Outpatient<br>Cost-Sharing Effect | -0.804***<br>(0.0952) | -2.765***<br>(0.162) | -0.793***<br>(0.110) | -3.219***<br>(0.183) |
| Observations | 5,220,007 | 5,220,007 | 2,996,596 | 2,996,596 |
| Bandwidth | 0.5 | 0.5 | 0.3 | 0.3 |
| Mean below threshold | 10.27 | 24.45 | 10.38 | 25.17 |
| Mean above threshold | 9.464 | 21.68 | 9.584 | 21.95 |
| Elasticity | -0.0685 | -0.101 | -0.0667 | -0.115 |

\*\*\* p<0.01, \*\* p<0.05, \* p<0.1

**Table S8.** Cumulative Effect of Outpatient Cost-Sharing on Chronic Disease Diagnosis (1-Year and 7-Year Horizons, at the 5 Monthly Minimum Wage (MMW) Threshold). Effect of greater outpatient cost-sharing on the probabilities (percentage points) of detecting chronic diseases for the same year and for all 7 years in the sample at the 5 Monthly Minimum Wage (MMW) Threshold. We use a sample of individuals with median monthly earnings within 0.5 and 0.3 MMWs of the threshold, and who worked during the entire year (12 months). We use ordinary least square with a second-order polynomial of the monthly earnings, and we compute heteroskedastic standard errors clustered at the individual level. We include a vector of time-invariant individual characteristics (age in 2011; sex; enrollment with a public (vs. private) insurer; and geographic region of enrollment, of which there are 5 in Colombia).

| Chronic Disease<br>Diagnosis | (1)<br>1 year | (2)<br>7 years | (3)<br>1 year | (4)<br>7 years |
| --- | --- | --- | --- | --- |
| Outpatient<br>Cost-Sharing Effect | 0.155<br>(0.156) | -0.368<br>(0.257) | -0.0260<br>(0.176) | -0.582**<br>(0.286) |
| Observations | 824,970 | 824,970 | 492,492 | 492,492 |
| Bandwidth | 0.5 | 0.5 | 0.3 | 0.3 |
| Mean below threshold | 9.324 | 22.36 | 9.299 | 22.28 |
| Mean above threshold | 9.479 | 21.99 | 9.273 | 21.70 |
| Elasticity | 0.0183 | -0.0184 | -0.00311 | -0.0294 |

\*\*\* p<0.01, \*\* p<0.05, \* p<0.1

**Table S9.** Cumulative Effect of Outpatient Cost-Sharing on Potentially-Avoidable Hospital Service Use (0.5 Monthly Minimum Wage (MMW) Bandwidth, at the 2 MMW Threshold). Effect of greater outpatient cost-sharing on the use of potentially avoidable hospital services – including inpatient stays, emergency room visits, and use of intensive care units at the 2 Monthly Minimum Wage (MMW) Threshold. We use a sample of individuals with median monthly earnings within 0.5 MMWs of the threshold, and who worked during the entire year (12 months). We use ordinary least square with a second-order polynomial of the monthly earnings, and we compute heteroskedastic standard errors clustered at the individual level. We include a vector of time-invariant individual characteristics (age in 2011; sex; enrollment with a public (vs. private) insurer; and geographic region of enrollment, of which there are 5 in Colombia). ER: is the probability (in percent points) to visit an ER in a calendar year; NUMER: is the number of visits to ER in a calendar year; HOS: is the probability (in percent points) to being hospitalized in a calendar year; NUMHOS: is the number of hospitalizations in a calendar year; NUMHOSDAY: is the number of days of hospital stay in a calendar year; ICU: is the probability (in percent points) to being hospitalized in an ICU in a calendar year; NUMICU: is the number of hospitalizations in ICU in a calendar year; NUMICUDAY: is the number of days of hospital stay in ICU in a calendar year.

| Potentially-Avoidable<br>Hospital Service Use | (1)<br>ER | (2)<br>NUMER | (3)<br>HOS | (4)<br>NUMHOS | (5)<br>NUMHOSDAY | (6)<br>ICU | (7)<br>NUMICU | (8)<br>NUMICUDAY |
| --- | --- | --- | --- | --- | --- | --- | --- | --- |
| Outpatient<br>Cost-Sharing Effect | 1.333***<br>(0.120) | 0.0489***<br>(0.00345) | 0.908***<br>(0.0562) | 0.101***<br>(0.0103) | 0.0256***<br>(0.00144) | 0.164***<br>(0.0145) | 0.0205***<br>(0.00261) | 0.00266***<br>(0.000354) |
| Observations | 5,220,007 | 5,220,007 | 5,220,007 | 5,220,007 | 5,220,007 | 5,220,007 | 5,220,007 | 5,220,007 |
| Mean below threshold | 27.10 | 0.530 | 4.572 | 0.171 | 0.0793 | 0.246 | 0.0195 | 0.00363 |
| Mean above threshold | 28.43 | 0.579 | 5.481 | 0.273 | 0.105 | 0.410 | 0.0400 | 0.00629 |
| Elasticity | 0.0403 | 0.0741 | 0.152 | 0.383 | 0.234 | 0.420 | 0.578 | 0.451 |

\*\*\* p<0.01, \*\* p<0.05, \* p<0.1

**Table S10.** Cumulative Effect of Outpatient Cost-Sharing on Potentially-Avoidable Hospital Service Use (0.3 Monthly Minimum Wage (MMW) Bandwidth, at the 2 MMW Threshold). Effect of greater outpatient cost-sharing on the use of potentially avoidable hospital services – including inpatient stays, emergency room visits, and use of intensive care units at the 2 Monthly Minimum Wage (MMW) Threshold. We use a sample of individuals with median monthly earnings within 0.3 MMWs of the threshold, and who worked during the entire year (12 months). We use ordinary least square with a second-order polynomial of the monthly earnings, and we compute heteroskedastic standard errors clustered at the individual level. We include a vector of time-invariant individual characteristics (age in 2011; sex; enrollment with a public (vs. private) insurer; and geographic region of enrollment, of which there are 5 in Colombia). ER: is the probability (in percent points) to visit an ER in a calendar year; NUMER: is the number of visits to ER in a calendar year; HOS: is the probability (in percent points) to being hospitalized in a calendar year; NUMHOS: is the number of hospitalizations in a calendar year; NUMHOSDAY: is the number of days of hospital stay in a calendar year; ICU: is the probability (in percent points) to being hospitalized in an ICU in a calendar year; NUMICU: is the number of hospitalizations in ICU in a calendar year; NUMICUDAY: is the number of days of hospital stay in ICU in a calendar year.

| Potentially-Avoidable<br>Hospital Service Use | (1)<br>ER | (2)<br>NUMER | (3)<br>HOS | (4)<br>NUMHOS | (5)<br>NUMHOSDAY | (6)<br>ICU | (7)<br>NUMICU | (8)<br>NUMICUDAY |
| --- | --- | --- | --- | --- | --- | --- | --- | --- |
| Outpatient<br>Cost-Sharing Effect | 1.830***<br>(0.141) | 0.0687***<br>(0.00404) | 1.259***<br>(0.0678) | 0.117***<br>(0.0138) | 0.0333***<br>(0.00176) | 0.207***<br>(0.0181) | 0.0217***<br>(0.00315) | 0.00287***<br>(0.000444) |
| Observations | 2,996,596 | 2,996,596 | 2,996,596 | 2,996,596 | 2,996,596 | 2,996,596 | 2,996,596 | 2,996,596 |
| Mean below threshold | 26.65 | 0.513 | 4.364 | 0.159 | 0.0742 | 0.218 | 0.0175 | 0.00337 |
| Mean above threshold | 28.48 | 0.582 | 5.624 | 0.276 | 0.108 | 0.425 | 0.0391 | 0.00624 |
| Elasticity | 0.0558 | 0.105 | 0.212 | 0.453 | 0.308 | 0.541 | 0.642 | 0.502 |

\*\*\* p<0.01, \*\* p<0.05, \* p<0.1

**Table S11.**
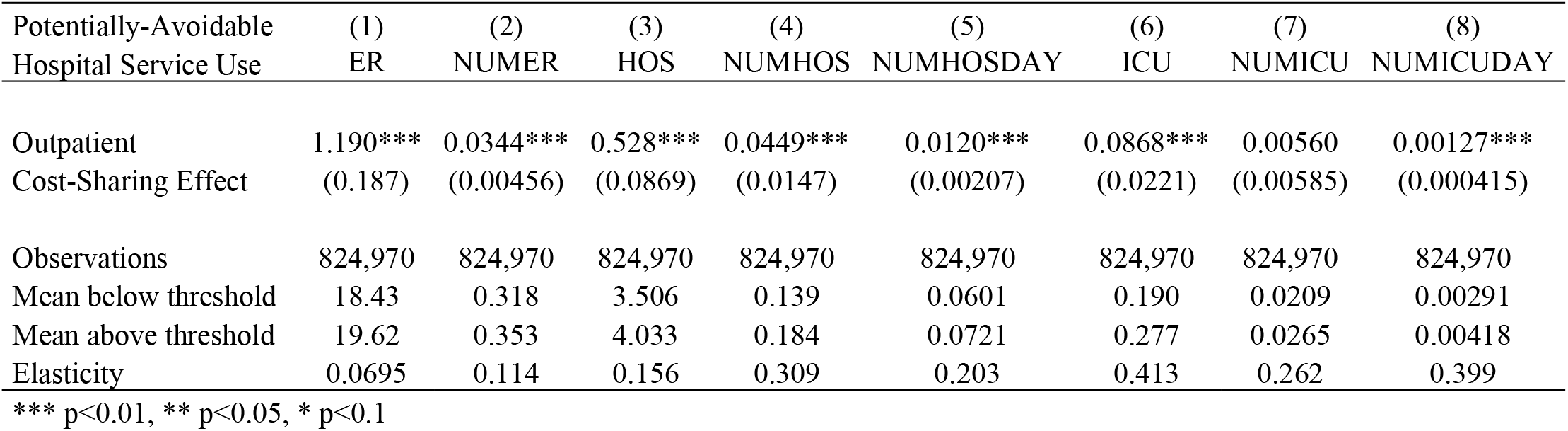
Cumulative Effect of Outpatient Cost-Sharing on Potentially-Avoidable Hospital Service Use (0.5 Monthly Minimum Wage (MMW) Bandwidth, at the 5 MMW Threshold). Effect of greater outpatient cost-sharing on the use of potentially avoidable hospital services – including inpatient stays, emergency room visits, and use of intensive care units at the 5 Monthly Minimum Wage (MMW) Threshold. We use a sample of individuals with median monthly earnings within 0.5 MMWs of the threshold, and who worked during the entire year (12 months). We use ordinary least square with a second-order polynomial of the monthly earnings, and we compute heteroskedastic standard errors clustered at the individual level. We include a vector of time-invariant individual characteristics (age in 2011; sex; enrollment with a public (vs. private) insurer; and geographic region of enrollment, of which there are 5 in Colombia). ER: is the probability (in percent points) to visit an ER in a calendar year; NUMER: is the number of visits to ER in a calendar year; HOS: is the probability (in percent points) to being hospitalized in a calendar year; NUMHOS: is the number of hospitalizations in a calendar year; NUMHOSDAY: is the number of days of hospital stay in a calendar year; ICU: is the probability (in percent points) to being hospitalized in an ICU in a calendar year; NUMICU: is the number of hospitalizations in ICU in a calendar year; NUMICUDAY: is the number of days of hospital stay in ICU in a calendar year.

**Table S12.** Cumulative Effect of Outpatient Cost-Sharing on Potentially-Avoidable Hospital Service Use (0.3 Monthly Minimum Wage (MMW) Bandwidth, at the 5 MMW Threshold). Effect of greater outpatient cost-sharing on the use of potentially avoidable hospital services – including inpatient stays, emergency room visits, and use of intensive care units at the 5 Monthly Minimum Wage (MMW) Threshold. We use a sample of individuals with median monthly earnings within 0.3 MMWs of the threshold, and who worked during the entire year (12 months). We use ordinary least square with a second-order polynomial of the monthly earnings, and we compute heteroskedastic standard errors clustered at the individual level. We include a vector of time-invariant individual characteristics (age in 2011; sex; enrollment with a public (vs. private) insurer; and geographic region of enrollment, of which there are 5 in Colombia). ER: is the probability (in percent points) to visit an ER in a calendar year; NUMER: is the number of visits to ER in a calendar year; HOS: is the probability (in percent points) to being hospitalized in a calendar year; NUMHOS: is the number of hospitalizations in a calendar year; NUMHOSDAY: is the number of days of hospital stay in a calendar year; ICU: is the probability (in percent points) to being hospitalized in an ICU in a calendar year; NUMICU: is the number of hospitalizations in ICU in a calendar year; NUMICUDAY: is the number of days of hospital stay in ICU in a calendar year.

| Potentially-Avoidable<br>Hospital Service Use | (1)<br>ER | (2)<br>NUMER | (3)<br>HOS | (4)<br>NUMHOS | (5)<br>NUMHOSDAY | (6)<br>ICU | (7)<br>NUMICU | (8)<br>NUMICUDAY |
| --- | --- | --- | --- | --- | --- | --- | --- | --- |
| Outpatient<br>Cost-Sharing Effect | 0.737***<br>(0.213) | 0.0229***<br>(0.00512) | 0.369***<br>(0.100) | 0.0349*<br>(0.0183) | 0.00960***<br>(0.00236) | 0.0821***<br>(0.0254) | 0.00443<br>(0.00743) | 0.00129***<br>(0.000479) |
| Observations | 492,492 | 492,492 | 492,492 | 492,492 | 492,492 | 492,492 | 492,492 | 492,492 |
| Mean below threshold | 18.61 | 0.323 | 3.553 | 0.143 | 0.0603 | 0.186 | 0.0218 | 0.00278 |
| Mean above threshold | 19.35 | 0.346 | 3.922 | 0.178 | 0.0699 | 0.268 | 0.0262 | 0.00407 |
| Elasticity | 0.0432 | 0.0760 | 0.110 | 0.241 | 0.164 | 0.402 | 0.205 | 0.419 |

\*\*\* p<0.01, \*\* p<0.05, \* p<0.1

**Table S13.**
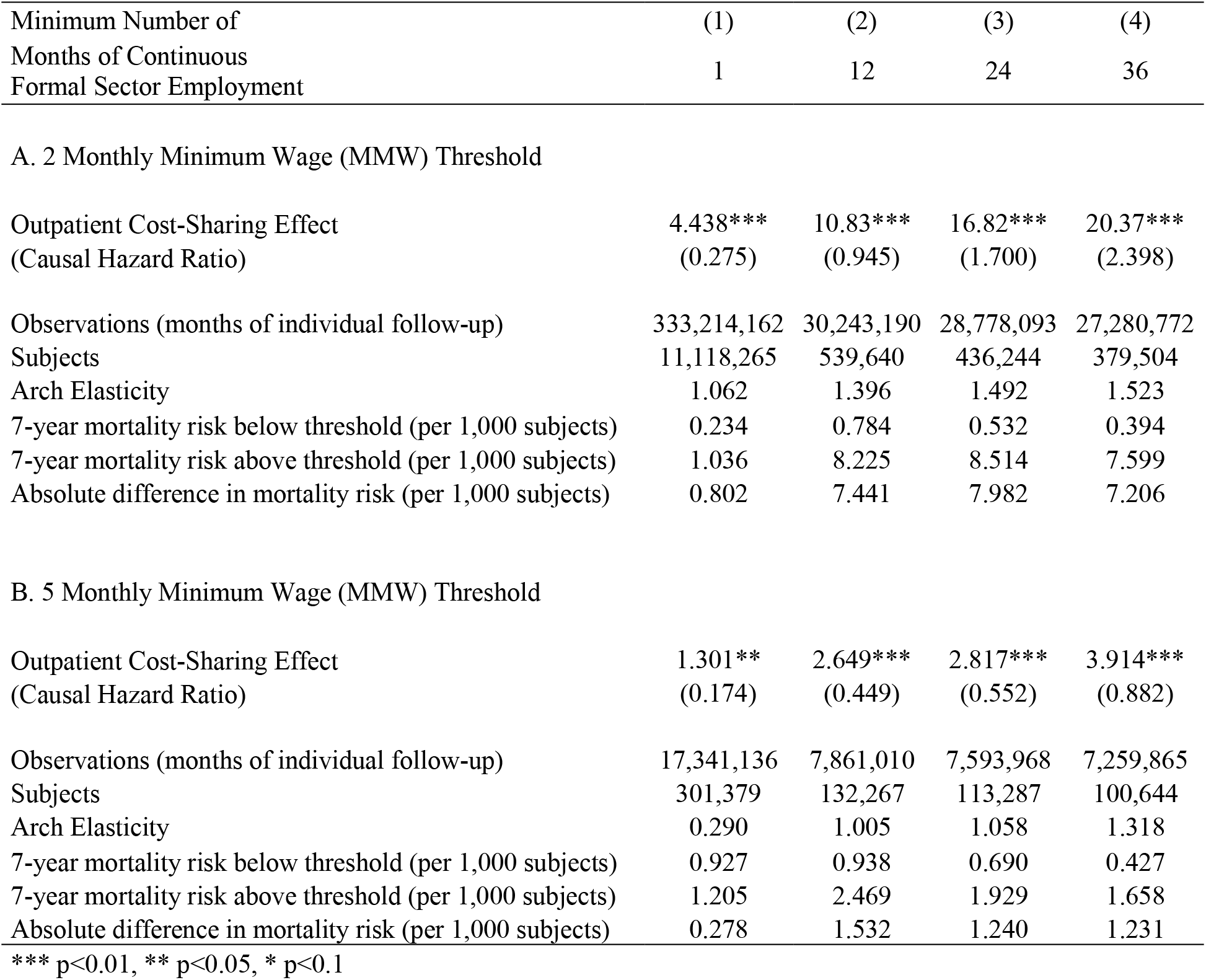
Cumulative Effect of Outpatient Cost-Sharing on 7-Year Mortality Hazard (Time-Dependent Cox Models, at the 2 and 5 Monthly Minimum Wage (MMW) Thresholds). Outpatient cost-sharing hazard ratios for 7-year mortality hazard at both thresholds (2 MMW **(A)** and 5 MMW **(B)**) using time-dependent Cox models among samples of individuals continuously employed in the formal sector for varying amounts of time (1, 12, 24 and 36 months), and with median monthly earnings within 1 MMW of the thresholds. We use time-dependent Cox models with a second-order polynomial of the monthly earnings. We include a vector of time-invariant individual characteristics (age in 2011; sex; enrollment with a public (vs. private) insurer; and geographic region of enrollment, of which there are 5 in Colombia).

| Minimum Number of<br>Months of Continuous<br>Formal Sector Employment | (1) | (2) | (3) | (4) |
| --- | --- | --- | --- | --- |
|  | 1 | 12 | 24 | 36 |
| **A. 2 Monthly Minimum Wage (MMW) Threshold** |  |  |  |  |
| Outpatient Cost-Sharing Effect<br>(Causal Hazard Ratio) | 4.438***<br>(0.275) | 10.83***<br>(0.945) | 16.82***<br>(1.700) | 20.37***<br>(2.398) |
| Observations (months of individual follow-up) | 333,214,162 | 30,243,190 | 28,778,093 | 27,280,772 |
| Subjects | 11,118,265 | 539,640 | 436,244 | 379,504 |
| Arch Elasticity | 1.062 | 1.396 | 1.492 | 1.523 |
| 7-year mortality risk below threshold (per 1,000 subjects) | 0.234 | 0.784 | 0.532 | 0.394 |
| 7-year mortality risk above threshold (per 1,000 subjects) | 1.036 | 8.225 | 8.514 | 7.599 |
| Absolute difference in mortality risk (per 1,000 subjects) | 0.802 | 7.441 | 7.982 | 7.206 |

**Table S13. Cumulative Effect of Outpatient Cost-Sharing on 7-Year Mortality Hazard (Time-Dependent Cox Models, at the 2 and 5 Monthly Minimum Wage (MMW) Thresholds).**
|  |  |  |  |  |
| --- | --- | --- | --- | --- |
| Outpatient Cost-Sharing Effect<br>(Causal Hazard Ratio) | 1.301**<br>(0.174) | 2.649***<br>(0.449) | 2.817***<br>(0.552) | 3.914***<br>(0.882) |
| Observations (months of individual follow-up) | 17,341,136 | 7,861,010 | 7,593,968 | 7,259,865 |
| Subjects | 301,379 | 132,267 | 113,287 | 100,644 |
| Arch Elasticity | 0.290 | 1.005 | 1.058 | 1.318 |
| 7-year mortality risk below threshold (per 1,000 subjects) | 0.927 | 0.938 | 0.690 | 0.427 |
| 7-year mortality risk above threshold (per 1,000 subjects) | 1.205 | 2.469 | 1.929 | 1.658 |
| Absolute difference in mortality risk (per 1,000 subjects) | 0.278 | 1.532 | 1.240 | 1.231 |

\*\*\* p<0.01, \*\* p<0.05, \* p<0.1

**Table S14.**
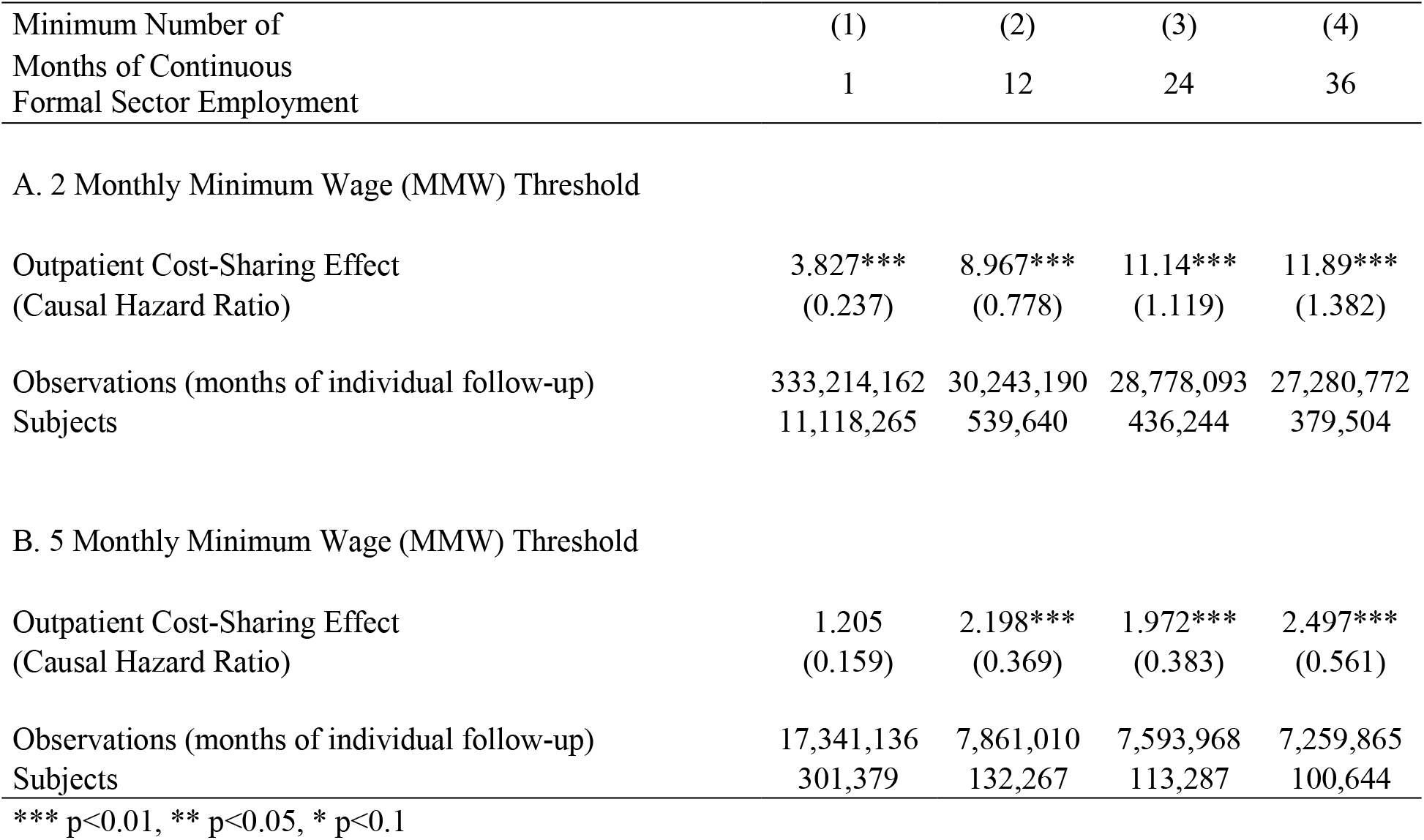
Cumulative Effect of Outpatient Cost-Sharing on 7-Year Mortality Hazard (Parametric survival model (Exponential Distribution), at the 2 and 5 Monthly Minimum Wage (MMW) Thresholds). Outpatient cost-sharing hazard ratios for 7-year mortality hazard at both thresholds (2 MMW **(A)** and 5 MMW **(B)**) using time-dependent parametric survival model with an Exponential Distribution among samples of individuals continuously employed in the formal sector for varying amounts of time (1, 12, 24 and 36 months), and with median monthly earnings within 1 MMW of the thresholds. We use a second-order polynomial of the monthly earnings. We include a vector of time-invariant individual characteristics (age in 2011; sex; enrollment with a public (vs. private) insurer; and geographic region of enrollment, of which there are 5 in Colombia).

| Minimum Number of<br>Months of Continuous<br>Formal Sector Employment | (1) | (2) | (3) | (4) |
| --- | --- | --- | --- | --- |
|  | 1 | 12 | 24 | 36 |
| A. 2 Monthly Minimum Wage (MMW) Threshold |  |  |  |  |
| Outpatient Cost-Sharing Effect<br>(Causal Hazard Ratio) | 3.827***<br>(0.237) | 8.967***<br>(0.778) | 11.14***<br>(1.119) | 11.89***<br>(1.382) |
| Observations (months of individual follow-up) | 333,214,162 | 30,243,190 | 28,778,093 | 27,280,772 |
| Subjects | 11,118,265 | 539,640 | 436,244 | 379,504 |

**Table S14. Cumulative Effect of Outpatient Cost-Sharing on 7-Year Mortality Hazard (Parametric survival model (Exponential Distribution), at the 2 and 5 Monthly Minimum Wage (MMW) Thresholds).**
|  |  |  |  |  |
| --- | --- | --- | --- | --- |
| Outpatient Cost-Sharing Effect<br>(Causal Hazard Ratio) | 1.205<br>(0.159) | 2.198***<br>(0.369) | 1.972***<br>(0.383) | 2.497***<br>(0.561) |
| Observations (months of individual follow-up) | 17,341,136 | 7,861,010 | 7,593,968 | 7,259,865 |
| Subjects | 301,379 | 132,267 | 113,287 | 100,644 |

\*\*\* p<0.01, \*\* p<0.05, \* p<0.1

**Table S15.**
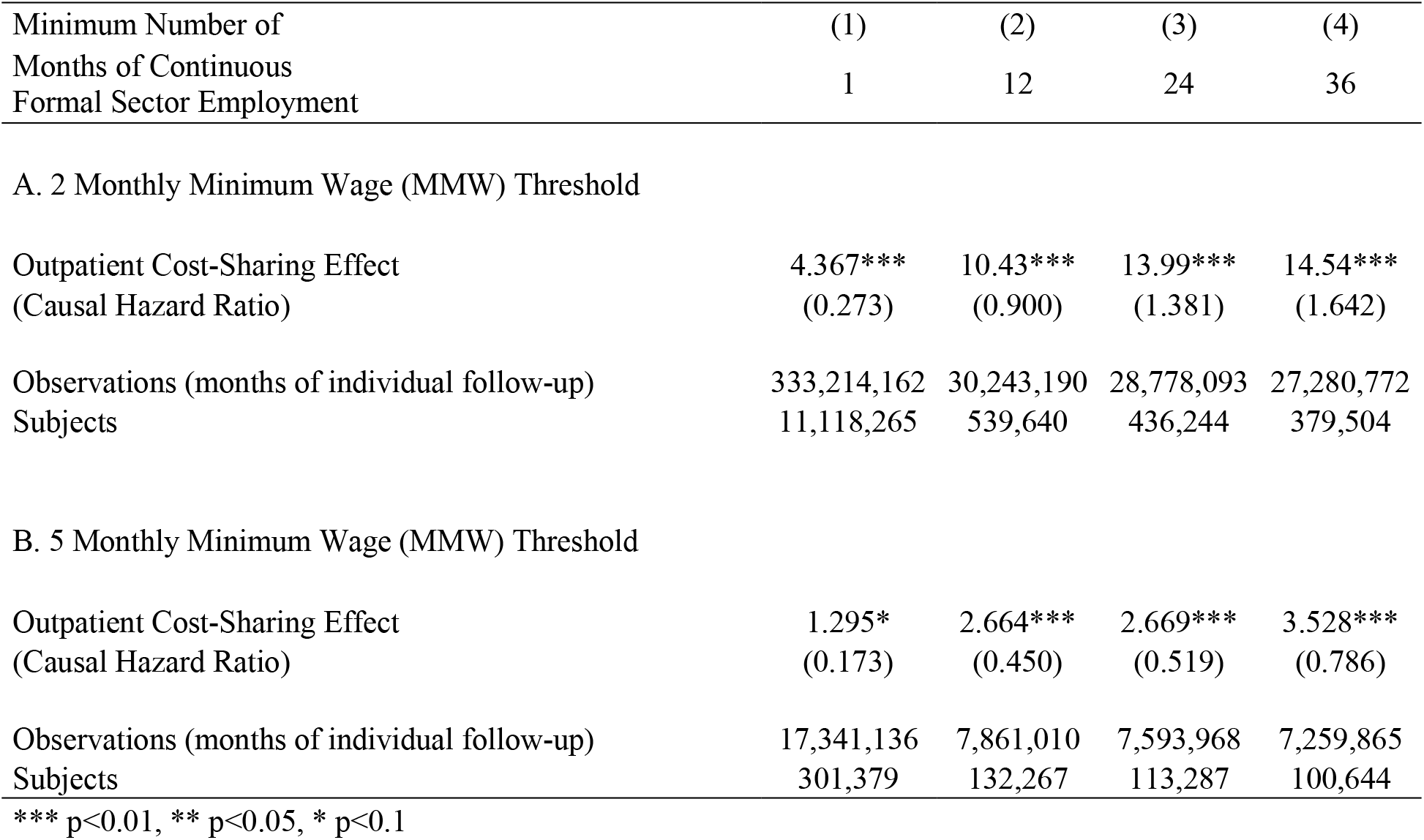
Cumulative Effect of Outpatient Cost-Sharing on 7-Year Mortality Hazard (Parametric survival model (Weibull Distribution), at the 2 and 5 Monthly Minimum Wage (MMW) Thresholds). Outpatient cost-sharing hazard ratios for 7-year mortality hazard at both thresholds (2 MMW **(A)** and 5 MMW **(B)**) using time-dependent parametric survival model with a Weibull Distribution among samples of individuals continuously employed in the formal sector for varying amounts of time (1, 12, 24 and 36 months), and with median monthly earnings within 1 MMW of the thresholds. We use a second-order polynomial of the monthly earnings. We include a vector of time-invariant individual characteristics (age in 2011; sex; enrollment with a public (vs. private) insurer; and geographic region of enrollment, of which there are 5 in Colombia).

| Minimum Number of<br>Months of Continuous<br>Formal Sector Employment | (1) | (2) | (3) | (4) |
| --- | --- | --- | --- | --- |
|  | 1 | 12 | 24 | 36 |
| A. 2 Monthly Minimum Wage (MMW) Threshold |  |  |  |  |
| Outpatient Cost-Sharing Effect<br>(Causal Hazard Ratio) | 4.367***<br>(0.273) | 10.43***<br>(0.900) | 13.99***<br>(1.381) | 14.54***<br>(1.642) |
| Observations (months of individual follow-up) | 333,214,162 | 30,243,190 | 28,778,093 | 27,280,772 |
| Subjects | 11,118,265 | 539,640 | 436,244 | 379,504 |

**Table S15. Cumulative Effect of Outpatient Cost-Sharing on 7-Year Mortality Hazard (Parametric survival model (Weibull Distribution), at the 2 and 5 Monthly Minimum Wage (MMW) Thresholds).**
|  |  |  |  |  |
| --- | --- | --- | --- | --- |
| Outpatient Cost-Sharing Effect<br>(Causal Hazard Ratio) | 1.295*<br>(0.173) | 2.664***<br>(0.450) | 2.669***<br>(0.519) | 3.528***<br>(0.786) |
| Observations (months of individual follow-up) | 17,341,136 | 7,861,010 | 7,593,968 | 7,259,865 |
| Subjects | 301,379 | 132,267 | 113,287 | 100,644 |

\*\*\* p<0.01, \*\* p<0.05, \* p<0.1

## Footnotes

1 A recent working paper shows that higher cost-sharing for drugs increases mortality among those 65 years of age in the United States (Chandra, Flack, and Obermeyer 2021). For a systematic review of the effect of user fees and health insurance on health outcomes in lower-income countries, see Qin et al. (2019).

2 The price elasticities of services related to the detection and long-term management of major chronic diseases may also be larger (i.e., service use may be more sensitive to prices) than for services addressing acute illnesses. Recent developments in value-based insurance design could, in principle, help to structure patient cost sharing to differentially encourage use of higher vs. lower value health services, but the data requirements are onerous, and potentially infeasible in many countries (Chernew, Rosen, and Fendrick 2007; Goldman, Joyce, and Karaca-Mandic 2006; Rosen et al. 2005).

3 Previous studies have shown that health insurance can lead to reductions in mortality (Sommers, Baicker, and Epstein 2012; Sommers, Long, and Baicker 2014; Goldin, Lurie, and McCubbin 2021; Sood et al. 2014; Card, Dobkin, and Maestas 2009; Miller, Johnson, and Wherry 2019). However, health insurance bundles together a variety of differences in the availability and quality of medical care as well as incentives on the supply-side – hence studying the consequences of health insurance is conceptually distinct from our specific focus on demand-side cost sharing.

4 The annual service use results shown in Figure 2 are estimated using the general equation shown in the following paragraph.

5 An elasticity is the ratio of the percent change in quantity of services to the percent change in cost-sharing (or price). The larger the absolute value of the elasticity, the more sensitive the service is to cost-sharing.

6 Supplement Tables S5 and S6.

1 *w*_*imt*_ *=* W_*imt*_/*MMW*_*t*_ where the numerator is individual’s *i* earnings (in Colombian Pesos) in month *m* and year *t*, and *MMW*_*t*_ is the legal monthly minimum wage in year *t*.

